# LSD use in the United States: Examining user demographics and their evolution from 2015-2019

**DOI:** 10.1101/2022.12.23.22283883

**Authors:** Jeremy Weleff, Akhil Anand, Elizabeth N. Dewey, Brian S. Barnett

**Author notes:** **Corresponding author*** Jeremy Weleff, DO, Yale University School of Medicine / Cleveland Clinic, Neurological Institute, Department of Psychiatry and Psychology, 9500 Euclid Ave, Cleveland, Ohio 44195, Twitter Jeremy Weleff: @JeremyWeleff, Twitter Brian Barnett: @BrianBarnettMD. **Disclosure Statement** Dr. Barnett has received stock options from CB Therapeutics as compensation for advisory services and monetary compensation for editorial work for DynaMed Plus (EBSCO Industries, Inc) and consulting services for Cerebral. The other authors report no potential conflicts of interest. **Funding** This work was unfunded. **Data availability statement** Data are publicly available. **Author Contributions: Jeremy Weleff:** Conceptualization, Methodology, Writing - Original Draft Writing, Writing - Review and Editing **Elizabeth Dewey:** Data curation, Methodology, Formal Analysis **Akhil Anand:** Conceptualization, Writing - Review and Editing **Brian Barnett:** Conceptualization, Methodology, Writing - Original Draft Writing, Writing - Review and Editing, Supervision.

## Abstract

**Background:** Lysergic acid (LSD) use has risen in the United States (US) in recent years amid increased interest in therapeutic applications of psychedelics. Despite this, contemporary epidemiological investigations of LSD users are few. To expand the literature on this topic, we sought to characterize past-year LSD users in the US and investigate recent demographic evolution within this population.

**Methods:** Using National Survey on Drug Use and Health (NSDUH) data from 2015-2019, we investigated correlates of past-year LSD use and associated changes over the study period.

**Results:** Past-year LSD use increased by 47% from 2015 to 2019 (0.59% to 0.87%). However, among people reporting past-year hallucinogen use disorder there was no significant proportional increase in LSD users. Notable correlates of LSD use on multivariable analysis were: increased LSD access, lower perceived risk of trying LSD, Asian race, low income, fewer children in the home, history of ever selling drugs, being approached by someone selling drugs in the past month, lack of influence of religious beliefs on decision-making, and past-year suicide attempt among people age 18 and older. We found no associations with unemployment, arrest history, past-year psychological distress, or sexually transmitted infections. From 2015 to 2019, the proportion of respondents reporting past-year LSD use who were pregnant, age 26-34, and married increased. Past-year LSD use among lifetime users of methamphetamine also rose.

**Conclusions:** Though still uncommonly used in the US, LSD’s societal acceptance may be increasing. Overall, LSD does not appear to contribute significantly to the country’s public health problems.

**Highlights:**

- For the last few years, NSDUH data has shown an increase in LSD use
- Despite this, rates of reported hallucinogen use disorder have not proportionally increased
- Evidence shows increasing societal acceptance for LSD use
- The proportion of respondents reporting past-year LSD use who were pregnant, age 26-34, and married increased
- Past-year LSD use among lifetime users of methamphetamine also rose

## Introduction

In 2019, there were estimated to be more than 27 million lifetime lysergic acid diethylamide (LSD) users in the United States (U.S.) (1). Use of LSD in the U.S. has more than doubled since the early 2000s (2) and past-year use remains on an upward trajectory (3). The factors behind this rise are not yet clear, though we speculate that the resurgence of interest in therapeutic applications of psychedelics and associated media may be possible contributors.

Early studies of LSD demonstrated therapeutic promise for its use in alcohol and opioid use disorders, as well as psychological distress associated with cancer (4–6). However, once LSD possession became illegal in the U.S. in 1971, clinical use and human subjects research assessing its therapeutic potential largely ceased. However, research into therapeutic applications of LSD and other psychedelics has returned to the U.S. in recent decades, despite a lack of federal funding to support this work (7). Social acceptability of use of LSD and other psychedelics may be growing, particularly around “wellness” applications of psychedelics. Although most psychedelics remain Schedule I substances in the U.S., a small but growing number of jurisdictions throughout the country have decriminalized or legalized possession of psychedelics. (8).

Despite sustained growth in LSD use, there have been few recent studies of LSD user demographics (2,3,9,10), leaving important gaps in our understanding of who is using LSD and how users might be changing. As a result, we sought to characterize past-year LSD users in the United States from 2015 to 2019 and evaluate whether user demography is evolving.

## Methods

### Data Description

The U.S. National Survey on Drug Use and Health (NSDUH) is conducted annually by the Substance Abuse and Mental Health Services Administration (SAMHSA), in all 50 states and the District of Columbia (11). NSDUH personnel administer the survey in person to randomly selected, noninstitutionalized civilians (important excluded populations include people who are imprisoned, hospitalized, or living in nursing homes). The survey inquires about substance use, mental health, other health-related issues, and treatments received for mental health conditions and substance use disorders. It also employs a sample-weighted design that includes weight adjustments for demographics, non-response, and other factors (12).

This study uses pooled data from NSDUH survey years 2015 to 2019 on all respondents (age 12 and older). Although 2020 survey data were available, data collection that year was disrupted due to COVID-19, and NSDUH administrators have advised researchers to avoid comparing 2020 data to those from previous years due to methodological alterations (13). The primary outcome for this study was past-year LSD use, which is an imputed variable in the NSDUH data set. There were no missing data for the outcome.

### Statistical Analysis

Unweighted frequencies for the primary outcome variable are reported, but unweighted frequencies for individual variables are not reported, similar to other publications that utilize NSDUH data. Continuous factors are summarized as weighted medians and interquartile ranges, and comparisons between LSD use groups were evaluated with log-transformed linear regression. Categorical factors are summarized using weighted percentages and 95% confidence intervals, and comparisons between LSD use groups were evaluated using Rao-Scott chi-square tests.

Univariable associations with LSD use were adjusted for survey year (2015-2019), age group (12-17, 18-25, 26-34, 35-49, 50+), gender (male, female), education (high school but no diploma, high school diploma/GED, some college/no degree, 2-year degree, 4-year degree), employment status (full-time, part-time, unemployed, other), marital status (never been married, married, divorced, widowed), ethnicity/race (White, Black, Asian, Hispanic, other), metropolitan area (small, large, non), religious beliefs (combination of importance and influences decision making), and criminal arrest history (yes/no). Associations were evaluated within each study year, and across study years for consistency. Where the association with LSD use was not consistent across survey years, orthogonal polynomial contrasts were used to determine if a linear trend was present.

Logistic regression was used to calculate adjusted odds ratios for the risk of past-year LSD use. All odds ratios were adjusted for the factors listed above. In addition to evaluating factors individually, three separate multivariable models were constructed: 1) for respondents under the age of 18; 2) for respondents over the age of 18; 3) for respondents of all ages. In the all ages model, questions that were not asked to respondents under the age of 18 or had answers that were nearly universal were recoded to either incorporate ages 12-17 as its own group or combined into the appropriate group (e.g., employment status). The model for respondents over age 18 included questions about mental health that could not be included in the all ages or under age 18 model. Multivariable models were constructed with factors associated with LSD use, after adjusting for the factors listed above. Models were simplified wherever appropriate to reduce complexity and to address multicollinearity and confounding.

Independent variables in our multivariable logistic regression models consisted primarily of factors previously associated with LSD use. For details about variable selection, please see Appendix A of the supplement.

Survey analysis procedures were used for all analyses due to the complex survey design, utilizing the NSDUH respondent weight, replicate, and variance strata. Since this study utilizes five years of survey data, the respondent weight was divided by five for analyses that included all study years. Significance was assessed at *p*<0.05. Analysis was conducted in SAS 9.4 (SAS Inc., Cary, NC).

### Ethics

This study was approved by the Cleveland Clinic Institutional Review Board.

## Results

### Demographic Characteristics of LSD Users

The final unweighted sample size was N=282,768, and 3,632 respondents reported past-year LSD use (past-year weighted prevalence 0.77%, 95% Confidence Interval: 0.72-0.81%). All investigated respondent factors were significantly associated with past-year LSD use on univariate analysis except for number of religious services attended in the past year and participation in government assistance programs in the current year. Compared to non-LSD users, past-year LSD users were disproportionately: male (67.9% vs 48.3%), between the ages of 18-25 (56.4% vs 12.3%), White (71.1% vs 62.8%), lesbian, gay, or bisexual (18.0% vs 4.9%), living in a large metropolitan area (59.4% vs 55.9%), never married (85.7% vs 28.1%), without children younger than 18 in the home (92.8% vs 73.3%), unemployed or employed part-time (35.1% vs 17.3% in age 18+), lower income (less than $20,000/year: 25.4% vs 16.1%), and uninsured (85.4% vs 90.7%). LSD users were also disproportionately likely to strongly disagree that their religious views influenced their decisions (45.7% vs 17.3%), strongly disagree that their religious views were a very important part of their life (43.8% vs 17.5%), report experiencing past-year serious psychological distress: (36.3% vs 11.1%), and report attempting suicide in the past year (3.9% vs 0.54%) (all *p*<0.001). For further details see Table 1.

**Table 1.** Demographic Characteristics by Past-Year LSD Use from NSDUH Survey Respondents from 2015-2019.

|  |  | Past-year LSD use |  |  |
| --- | --- | --- | --- | --- |
|  | All Respondents | No Past-year Use | Past-year Use |  |
| Survey Question | (N=282,768) | (N=279,136) | (N=3,632) | p-value |
| Survey Year |  |  |  | **<0.001<sup>c</sup>** |
| 2015 | 19.7 (19.4, 20.0) | 19.7 (19.4, 20.1) | 15.2 (13.6, 16.7) |  |
| 2016 | 19.8 (19.5, 20.1) | 19.8 (19.6, 20.1) | 18.4 (16.2, 20.6) |  |
| 2017 | 20.0 (19.8, 20.3) | 20.0 (19.8, 20.3) | 21.2 (19.0, 23.3) |  |
| 2018 | 20.2 (19.9, 20.4) | 20.1 (19.9, 20.4) | 22.2 (19.7, 24.7) |  |
| 2019 | 20.3 (19.9, 20.6) | 20.2 (19.9, 20.6) | 23.1 (20.9, 25.3) |  |
| Gender Male | 48.5 (48.2, 48.8) | 48.3 (48.1, 48.6) | 67.9 (65.7, 70.0) | **<0.001<sup>c</sup>** |
| Under Age 18 Gender Male | 50.9 (50.4, 51.5) | 50.8 (50.3, 51.3) | 61.5 (56.8, 66.2) | **<0.001<sup>c</sup>** |
| Age 18+ Gender Male | 48.3 (47.9, 48.6) | 48.1 (47.8, 48.4) | 68.7 (66.6, 70.8) | **<0.001<sup>c</sup>** |
| Age Groups |  |  |  | **<0.001<sup>c</sup>** |
| Age: 12-17 | 9.2 (9.0, 9.3) | 9.1 (9.0, 9.3) | 11.7 (10.4, 13.0) |  |
| Age: 18-25 | 12.6 (12.5, 12.8) | 12.3 (12.1, 12.5) | 56.4 (54.1, 58.8) |  |
| Age: 26-34 | 14.5 (14.3, 14.7) | 14.4 (14.2, 14.7) | 22.7 (20.8, 24.6) |  |
| Age: 35-49 | 22.4 (22.1, 22.7) | 22.5 (22.3, 22.8) | 6.6 (5.3, 8.0) |  |
| Age: 50+ | 41.3 (40.8, 41.8) | 41.6 (41.1, 42.1) | 2.6 (1.2, 4.0) |  |
| Age Groups, Over/Under 18 |  |  |  | **<0.001<sup>c</sup>** |
| Under18 | 9.2 (9.0, 9.3) | 9.1 (9.0, 9.3) | 11.7 (10.4, 13.0) |  |
| 18+ Years | 90.8 (90.7, 91.0) | 90.9 (90.7, 91.0) | 88.3 (87.0, 89.6) |  |
| Race/Hispanicity* |  |  |  | **<0.001<sup>c</sup>** |
| Nonhisp White | 62.8 (62.3, 63.3) | 62.8 (62.3, 63.3) | 71.1 (68.7, 73.5) |  |

| Survey Question | All Respondents<br>(N=282,768) | Past-year LSD use |  | p-value |
| --- | --- | --- | --- | --- |
|  |  | No Past-year Use<br>(N=279,136) | Past-year Use<br>(N=3,632) |  |
| Non-Hispanic Black/African American | 12.0 (11.7, 12.4) | 12.1 (11.7, 12.4) | 4.5 (3.6, 5.3) |  |
| Non-Hispanic Native American/Alaskan Native | 0.56 (0.51, 0.61) | 0.56 (0.51, 0.61) | 0.44 (0.22, 0.66) |  |
| Non-Hispanic Native Hawaiian/Pacific Islander | 0.38 (0.33, 0.42) | 0.38 (0.33, 0.42) | 0.37 (0.16, 0.58) |  |
| Non-Hispanic Asian | 5.6 (5.4, 5.8) | 5.6 (5.4, 5.8) | 4.2 (3.1, 5.3) |  |
| Non-Hispanic More Than One Race | 1.8 (1.8, 1.9) | 1.8 (1.8, 1.9) | 3.9 (3.0, 4.7) |  |
| Hispanic | 16.8 (16.4, 17.2) | 16.8 (16.4, 17.2) | 15.6 (13.4, 17.7) |  |
| Sexual Identity, Age 18+ Only |  |  |  | **<0.001<sup>c</sup>** |
| Heterosexual | 94.3 (94.2, 94.5) | 94.4 (94.3, 94.6) | 81.3 (79.2, 83.3) |  |
| Lesbian Or Gay | 1.9 (1.8, 2.0) | 1.9 (1.8, 2.0) | 4.2 (3.3, 5.0) |  |
| Bisexual | 3.1 (3.0, 3.2) | 3.0 (2.9, 3.1) | 13.8 (12.2, 15.3) |  |
| Don't Know | 0.64 (0.58, 0.70) | 0.64 (0.58, 0.70) | 0.78 (0.28, 1.3) |  |
| Urbanicity |  |  |  | **<0.001<sup>c</sup>** |
| Large Metro | 55.9 (55.4, 56.4) | 55.9 (55.4, 56.4) | 59.4 (56.9, 61.8) |  |
| Small Metro | 29.9 (29.5, 30.4) | 29.9 (29.5, 30.4) | 29.3 (27.1, 31.5) |  |
| Nonmetro | 14.1 (13.8, 14.5) | 14.2 (13.8, 14.5) | 11.3 (9.7, 12.9) |  |
| Education* |  |  |  | **<0.001<sup>c</sup>** |
| Under Age 18 |  |  |  | **<0.001<sup>c</sup>** |
| Some HS, No Diploma | 98.9 (98.8, 99.0) | 99.0 (98.9, 99.1) | 94.9 (92.5, 97.4) |  |
| HS Diploma/GED | 0.87 (0.78, 0.96) | 0.83 (0.74, 0.91) | 4.9 (2.5, 7.3) |  |
| Some College, No Degree | 0.15 (0.11, 0.20) | 0.15 (0.11, 0.19) | 0.19 (0.00, 0.57) |  |
| 2-Year College Degree | 0.00 (0.00, 0.01) | 0.00 (0.00, 0.01) | -- |  |

| Survey Question | All Respondents<br>(N=282,768) | Past-year LSD use |  | p-value |
| --- | --- | --- | --- | --- |
|  |  | No Past-year Use<br>(N=279,136) | Past-year Use<br>(N=3,632) |  |
| 4-Year College Degree | 0.03 (0.01, 0.05) | 0.03 (0.01, 0.05) | -- |  |
| Age 18+ |  |  |  | **<0.001<sup>c</sup>** |
| Some HS, No Diploma | 12.7 (12.4, 13.0) | 12.7 (12.5, 13.0) | 8.3 (7.1, 9.6) |  |
| HS Diploma/GED | 24.8 (24.5, 25.1) | 24.9 (24.5, 25.2) | 23.4 (21.1, 25.8) |  |
| Some College, No Degree | 21.6 (21.3, 21.8) | 21.4 (21.2, 21.7) | 37.1 (34.4, 39.9) |  |
| 2-Year College Degree | 9.3 (9.1, 9.5) | 9.3 (9.1, 9.5) | 7.4 (6.2, 8.7) |  |
| 4-Year College Degree | 31.6 (31.1, 32.1) | 31.7 (31.2, 32.1) | 23.7 (21.2, 26.1) |  |
| Marital Status*, Age 18+ Only |  |  |  | **<0.001<sup>c</sup>** |
| Married | 51.7 (51.3, 52.2) | 52.1 (51.6, 52.5) | 8.6 (6.5, 10.6) |  |
| Widowed | 5.9 (5.7, 6.1) | 5.9 (5.8, 6.1) | 0.57 (0.00, 1.2) |  |
| Divorced Or Separated | 13.8 (13.6, 14.1) | 13.9 (13.6, 14.2) | 5.1 (3.6, 6.6) |  |
| Never Been Married | 28.5 (28.2, 28.9) | 28.1 (27.7, 28.4) | 85.7 (83.4, 88.1) |  |
| # Children < 18 in Home* |  |  |  | **<0.001<sup>c</sup>** |
| None | 73.8 (73.6, 74.1) | 73.7 (73.4, 73.9) | 92.8 (91.6, 94.1) |  |
| One | 10.9 (10.8, 11.0) | 10.9 (10.8, 11.1) | 4.3 (3.4, 5.2) |  |
| Two | 9.7 (9.6, 9.9) | 9.8 (9.6, 9.9) | 2.3 (1.4, 3.1) |  |
| Three Or More | 5.6 (5.4, 5.7) | 5.6 (5.5, 5.7) | 0.62 (0.22, 1.02) |  |
| Employment Status* |  |  |  |  |
| Under 18 |  |  |  | **<0.001<sup>c</sup>** |
| Employed Full Time | 4.6 (4.3, 4.8) | 4.5 (4.2, 4.8) | 8.8 (5.9, 11.8) |  |
| Employed Part Time | 24.1 (23.6, 24.7) | 24.0 (23.4, 24.6) | 33.1 (28.1, 38.0) |  |
| Unemployed | 10.8 (10.4, 11.2) | 10.7 (10.3, 11.1) | 16.3 (12.8, 19.8) |  |
| Other | 60.5 (59.8, 61.2) | 60.8 (60.1, 61.5) | 41.7 (36.4, 47.1) |  |

|  |  | Past-year LSD use |  |  |
| --- | --- | --- | --- | --- |
|  | All Respondents | No Past-year Use | Past-year Use |  |
| Survey Question | (N=282,768) | (N=279,136) | (N=3,632) | p-value |
| Age 18+ |  |  |  | **<0.001<sup>c</sup>** |
| Employed Full Time | 49.4 (49.1, 49.8) | 49.4 (49.0, 49.8) | 50.6 (47.8, 53.3) |  |
| Employed Part Time | 13.1 (12.9, 13.3) | 13.0 (12.8, 13.2) | 25.4 (23.2, 27.7) |  |
| Unemployed | 4.3 (4.2, 4.5) | 4.3 (4.1, 4.4) | 9.7 (8.3, 11.2) |  |
| Other | 33.2 (32.8, 33.5) | 33.3 (32.9, 33.7) | 14.3 (12.6, 15.9) |  |
| How many employers, age 18+ only / Past-year |  |  |  | **<0.001<sup>c</sup>** |
| One | 74.8 (74.4, 75.1) | 75.0 (74.7, 75.4) | 48.5 (44.9, 52.1) |  |
| Two | 18.3 (18.0, 18.6) | 18.1 (17.8, 18.4) | 31.6 (29.1, 34.0) |  |
| Three | 4.4 (4.3, 4.6) | 4.3 (4.2, 4.5) | 13.2 (11.4, 15.1) |  |
| Four or more | 2.5 (2.4, 2.6) | 2.5 (2.4, 2.6) | 6.7 (5.3, 8.1) |  |
| Total Family Income* |  |  |  | **<0.001<sup>c</sup>** |
| Less Than \$20,000 | 16.2 (15.9, 16.5) | 16.1 (15.8, 16.4) | 25.4 (23.3, 27.5) | |
| \$20,000-\$49,999 | 29.3 (28.9, 29.6) | 29.3 (28.9, 29.7) | 29.7 (27.5, 31.8) | |
| \$50,000-\$74,999 | 15.8 (15.6, 16.1) | 15.9 (15.6, 16.1) | 14.6 (12.9, 16.4) | |
| \$75,000 Or More | 38.7 (38.1, 39.2) | 38.8 (38.2, 39.3) | 30.3 (27.6, 33.0) | |
| # Days skipped work / Past Month, Median [Q1, Q3] | 0.00 [0.00, 0.00] | 0.00 [0.00, 0.00] | 0.00 [0.00, 0.25] | **<0.001<sup>b</sup>** |
| Working Cell Phone | 97.7 (97.7, 97.8) | 97.7 (97.6, 97.8) | 98.9 (98.3, 99.5) | **0.004<sup>c</sup>** |
| Poverty Level (% Of Us Census Poverty Threshold)* |  |  |  | **<0.001<sup>c</sup>** |
| Living In Poverty | 14.8 (14.5, 15.1) | 14.7 (14.4, 15.0) | 20.5 (19.0, 22.0) |  |
| Income ↑ 2X Fed Pov Thresh | 20.0 (19.7, 20.3) | 20.0 (19.7, 20.2) | 22.0 (20.0, 23.9) |  |

|  |  | Past-year LSD use |  |  |
| --- | --- | --- | --- | --- |
|  | All Respondents | No Past-year Use | Past-year Use |  |
| Survey Question | (N=282,768) | (N=279,136) | (N=3,632) | p-value |
| Income >2x Fed Pov Thresh | 65.2 (64.8, 65.7) | 65.3 (64.8, 65.8) | 57.6 (55.4, 59.7) |  |
| Participated in 1+ govt assist progr* | 18.6 (18.2, 18.9) | 18.6 (18.2, 18.9) | 17.3 (15.6, 19.0) | 0.13<sup>c</sup> |
| Covered by any health insurance* | 90.6 (90.4, 90.8) | 90.7 (90.5, 90.9) | 85.4 (83.6, 87.2) | <0.001<sup>c</sup> |
| Body Mass Index (BMI) (Age 18+ Only), Median [Q1, Q3]* | 27.3 [23.7, 31.7] | 27.3 [23.7, 31.7] | 23.7 [21.4, 27.4] | <0.001<sup>b</sup> |
| Pregnant, Female Only |  |  |  | 0.024<sup>c</sup> |
| Yes | 3.2 (3.1, 3.4) | 3.3 (3.1, 3.4) | 1.8 (0.66, 3.0) |  |
| No | 96.6 (96.5, 96.8) | 96.6 (96.5, 96.8) | 98.2 (97.0, 99.3) |  |
| Don't Know | 0.14 (0.11, 0.16) | 0.14 (0.11, 0.16) | 0.02 (0.00, 0.06) |  |
| Had sexually transmitted disease / Past Month |  |  |  | <0.001<sup>c</sup> |
| Yes | 1.9 (1.8, 2.0) | 1.9 (1.8, 2.0) | 6.9 (5.7, 8.0) |  |
| No | 97.9 (97.8, 98.0) | 98.0 (97.9, 98.0) | 92.7 (91.6, 93.9) |  |
| Don't Know | 0.16 (0.14, 0.18) | 0.16 (0.14, 0.18) | 0.41 (0.11, 0.72) |  |
| # Religious services past 12 months |  |  |  | 0.066<sup>c</sup> |
| 25 Or More | 68.1 (67.8, 68.3) | 68.0 (67.8, 68.3) | 69.8 (67.9, 71.7) |  |
| Less Than 25 | 31.9 (31.7, 32.2) | 32.0 (31.7, 32.2) | 30.2 (28.3, 32.1) |  |
| My religious beliefs influence my decisions |  |  |  | <0.001<sup>c</sup> |
| Strongly Disagree | 17.5 (17.2, 17.7) | 17.3 (17.0, 17.5) | 45.7 (43.4, 47.9) |  |
| Disagree | 15.7 (15.5, 15.9) | 15.7 (15.5, 15.9) | 23.6 (22.2, 24.9) |  |
| Agree | 35.1 (34.9, 35.4) | 35.2 (34.9, 35.5) | 23.4 (21.6, 25.2) |  |
| Strongly Agree | 31.7 (31.4, 31.9) | 31.8 (31.6, 32.1) | 7.3 (6.0, 8.7) |  |

| Survey Question | All Respondents<br>(N=282,768) | Past-year LSD use |  | p-value |
| --- | --- | --- | --- | --- |
|  |  | No Past-year Use<br>(N=279,136) | Past-year Use<br>(N=3,632) |  |
| My religious beliefs are very important |  |  |  | **<0.001<sup>c</sup>** |
| Strongly Disagree | 17.7 (17.4, 17.9) | 17.5 (17.2, 17.7) | 43.8 (41.2, 46.3) |  |
| Disagree | 13.0 (12.8, 13.1) | 12.9 (12.7, 13.1) | 22.3 (20.5, 24.1) |  |
| Agree | 33.6 (33.3, 33.8) | 33.6 (33.3, 33.9) | 23.9 (22.1, 25.7) |  |
| Strongly Agree | 35.8 (35.5, 36.1) | 36.0 (35.7, 36.3) | 10.1 (8.6, 11.7) |  |
| Stolen/tried to steal, worth > \$50 | 1.04 (1.00, 1.08) | 0.99 (0.95, 1.03) | 7.6 (6.5, 8.7) | **<0.001<sup>c</sup>** |
| Sold illegal drugs | 1.5 (1.5, 1.6) | 1.4 (1.3, 1.4) | 23.2 (21.0, 25.5) | **<0.001<sup>c</sup>** |
| Attacked someone w/intent to seriously hurt them | 1.2 (1.2, 1.3) | 1.2 (1.1, 1.2) | 5.5 (4.5, 6.4) | **<0.001<sup>c</sup>** |
| Ever arrested and booked for breaking the law | 15.6 (15.4, 15.9) | 15.5 (15.3, 15.7) | 33.7 (31.2, 36.2) | **<0.001<sup>c</sup>** |
| # Times arrested and booked past 12 months |  |  |  | **<0.001<sup>c</sup>** |
| None | 87.9 (87.5, 88.3) | 88.2 (87.8, 88.6) | 69.4 (65.0, 73.9) |  |
| Once | 9.0 (8.6, 9.4) | 8.8 (8.4, 9.1) | 22.0 (18.3, 25.6) |  |
| Twice | 2.1 (1.9, 2.3) | 2.1 (1.9, 2.3) | 6.0 (3.9, 8.1) |  |
| Three Or More | 0.95 (0.83, 1.07) | 0.92 (0.80, 1.05) | 2.6 (1.4, 3.9) |  |
| Overnight Stay as Inpnt in Hosp / Past-year | 9.5 (9.4, 9.7) | 9.6 (9.4, 9.8) | 8.1 (6.9, 9.2) | **0.018<sup>c</sup>** |
| Serious psych distress / Past-year | 11.3 (11.1, 11.5) | 11.1 (10.9, 11.3) | 36.3 (33.8, 38.8) | **<0.001<sup>c</sup>** |
| Attempted to kill self / Past-year | 0.56 (0.52, 0.60) | 0.54 (0.50, 0.58) | 3.9 (2.9, 4.9) | **<0.001<sup>c</sup>** |

Frequencies presented are unweighted counts. Unless otherwise noted, weighted proportions and 95% confidence intervals are reported. \*Imputed or recoded variable reported. P-values: a=linear regression; b=linear regression with log transformation; c=Rao-Scott chi-square test. SAS Survey Procedures used for all analyses.

All respondents who reported drug use were disproportionately represented among past-year LSD users; aside from tobacco, alcohol, marijuana, and cocaine, respondents who had not used LSD in the past year rarely had lifetime experience with other drugs (<10% non-LSD users reported using each of the other drugs assessed). Prevalence of past-year hallucinogen use disorder was disproportionately higher in past-year LSD users than non-users (6.6% vs 0.06%), as was history of ever using a needle to inject drugs (7.1% vs 1.6%). Past-year LSD users also disproportionately reported that LSD was very easy or fairly easy to obtain (53.0% vs 12.8%). See Supplemental Table 1 for further details.

Past-year LSD users disproportionately reported that there was no risk in: smoking marijuana once or twice a week (65.9% vs 18.5%), trying LSD once or twice (34.9% vs 3.2%), using LSD once or twice a week (12.0% vs 1.4%), trying heroin once or twice (2.3% vs 1.2%), using cocaine once or twice a week (1.3% vs 1.04%), or having 5 or more drinks once or twice a week (5.8% vs 3.1%) (all p<0.001). A notable exception was using heroin once or twice a week, which 0.83% of LSD users reported as no risk compared to 0.91% of non-users (p=0.002). See Supplemental Table 2 for further details.

### Demographic changes from 2014-2019

Past-year LSD use significantly increased by 47% from 2015 to 2019 (0.59% to 0.87%, p<0.0001) (Table 2). Yearly relative increases were higher between 2015-2016 (20.3%) and 2016-2017 (14.1%) but were modest between 2017-2018 (3.7%) and 2018-2019 (3.6%). There were strong within-year associations with past-year LSD use for nearly all respondent factors for each study year, p<0.001; exceptions were metropolitan area (large metro range: 53.2% to 60.9%, p=0.002 to 0.61) and female pregnancy (range 0.8% to 3%, p=0.03 to 0.78). These associations remained consistent across the study period with a few notable exceptions. Past-year LSD users were predominantly between the ages of 18-25 in every year, but the proportion of users aged 26-34 increased nearly every year from 16.3% in 2015 to 26.5% in 2019 (*p*<0.0001). The proportion of past-year LSD users who were married also increased over the study period from 5.7% in 2015 to 10.1% in 2019 (*p*<0.0001), as did the proportion who reported being pregnant at the time of NSDUH participation (though not necessarily at time of LSD use), which increased from 1.1% in 2015 to 2.1% in 2019 (*p*<0.0001).

**Table 2.** Demographic Characteristics per Study Year by Past-Year LSD Use from NSDUH Survey Respondents from 2015-2019.

|  | **2015** | **2016** | **2017** | **2018** | **2019** | **2015 vs 2019** |
| --- | --- | --- | --- | --- | --- | --- |
| **Demographic Characteristics** | **(N=57,146)** | **(N=56,897)** | **(N=56,276)** | **(N=56,313)** | **(N=56,136)** | **p-value** |
| **Past-year LSD Use** | 0.59<br>(0.53, 0.65) | 0.71<br>(0.61, 0.80) | 0.81<br>(0.71, 0.91) | 0.84<br>(0.73, 0.96) | 0.87<br>(0.79, 0.96) | **<.0001** |
| **Change from Previous Year** | -- | 0.12%<br>(20.3%) | 0.1%<br>(14.1%) | 0.03%<br>(3.7%) | 0.03%<br>(3.6%) |  |
| **Age groups** |  |  |  |  |  | **0.01** |
| No LSD Use |  |  |  |  |  |  |
| age: 12-17 | 9.3<br>(9.0, 9.5) | 9.2<br>(9.0, 9.4) | 9.1<br>(8.9, 9.4) | 9.1<br>(8.8, 9.4) | 9.0<br>(8.7, 9.3) |  |
| age: 18-25 | 12.7<br>(12.4, 13.1) | 12.5<br>(12.1, 12.9) | 12.2<br>(11.8, 12.7) | 12.1<br>(11.8, 12.4) | 11.9<br>(11.6, 12.2) |  |
| age: 26-34 | 14.3<br>(13.8, 14.8) | 14.4<br>(13.9, 14.8) | 14.5<br>(14.1, 14.9) | 14.5<br>(14.0, 15.0) | 14.5<br>(14.1, 15.0) |  |
| age: 35-49 | 22.7<br>(22.1, 23.3) | 22.7<br>(22.1, 23.2) | 22.5<br>(22.0, 23.1) | 22.5<br>(22.1, 22.9) | 22.3<br>(21.8, 22.7) |  |
| age: 50+ | 41.0<br>(40.1, 41.9) | 41.3<br>(40.4, 42.1) | 41.6<br>(40.7, 42.5) | 41.8<br>(41.1, 42.5) | 42.3<br>(41.4, 43.1) |  |
| Past-year LSD Use |  |  |  |  |  |  |
| age: 12-17 | 16.1<br>(12.9, 19.2) | 10.7<br>(7.9, 13.5) | 12.5<br>(10.0, 14.9) | 8.4<br>(6.6, 10.2) | 12.0<br>(9.4, 14.6) |  |
| age: 18-25 | 63.7<br>(58.6, 68.7) | 61.0<br>(54.5, 67.4) | 57.2<br>(51.2, 63.2) | 52.5<br>(47.8, 57.2) | 51.2<br>(45.9, 56.5) |  |
| age: 26-34 | 16.3<br>(11.6, 20.9) | 21.3<br>(15.0, 27.5) | 17.9<br>(12.5, 23.3) | 28.8<br>(23.0, 34.5) | 26.5<br>(21.5, 31.4) |  |
| age: 35-49 | 2.5<br>(0.50, 4.4) | 7.1<br>(3.4, 10.8) | 7.4<br>(4.5, 10.2) | 7.9<br>(4.3, 11.6) | 7.1<br>(4.5, 9.7) |  |

|  | 2015 | 2016 | 2017 | 2018 | 2019 | 2015 vs 2019 |
| --- | --- | --- | --- | --- | --- | --- |
| Demographic Characteristics | (N=57,146) | (N=56,897) | (N=56,276) | (N=56,313) | (N=56,136) | *p*-value |
| age: 50+ | 1.5<br>(0.00, 3.7) | 0 | 5.1<br>(0.96, 9.1) | 2.4<br>(0.00, 5.1) | 3.3<br>(0.60, 5.9) |  |
| Gender male |  |  |  |  |  | **0.90** |
| No LSD Use | 48.3<br>(47.7, 49.0) | 48.3<br>(47.7, 49.0) | 48.4<br>(47.8, 48.9) | 48.3<br>(47.7, 49.0) | 48.3<br>(47.7, 49.0) |  |
| Past-year LSD Use | 67.7<br>(62.9, 72.5) | 69.6<br>(64.3, 75.0) | 65.0<br>(60.1, 70.0) | 68.4<br>(63.4, 73.5) | 68.6<br>(64.1, 73.2) |  |
| Sexual identity |  |  |  |  |  | **<.0001** |
| No LSD Use |  |  |  |  |  |  |
| Heterosexual, i.e. straight | 95.1<br>(94.8, 95.3) | 95.1<br>(94.7, 95.4) | 94.5<br>(94.2, 94.8) | 94.1<br>(93.8, 94.4) | 93.6<br>(93.3, 93.9) |  |
| Lesbian or Gay | 1.8<br>(1.6, 2.0) | 1.8<br>(1.6, 2.0) | 2.0<br>(1.7, 2.2) | 1.9<br>(1.7, 2.1) | 2.0<br>(1.8, 2.2) |  |
| Bisexual | 2.5<br>(2.4, 2.7) | 2.5<br>(2.2, 2.7) | 3.0<br>(2.8, 3.2) | 3.3<br>(3.1, 3.5) | 3.8<br>(3.6, 4.0) |  |
| Don't know | 0.60<br>(0.49, 0.72) | 0.65<br>(0.53, 0.78) | 0.57<br>(0.44, 0.70) | 0.71<br>(0.58, 0.84) | 0.66<br>(0.54, 0.78) |  |
| Past-year LSD Use |  |  |  |  |  |  |
| Heterosexual, i.e. straight | 85.1<br>(80.0, 90.2) | 82.2<br>(77.4, 86.9) | 80.7<br>(77.0, 84.5) | 83.3<br>(80.0, 86.7) | 76.5<br>(70.7, 82.3) |  |
| Lesbian or Gay | 3.4<br>(1.4, 5.3) | 6.4<br>(3.2, 9.7) | 2.9<br>(1.5, 4.2) | 3.4<br>(1.2, 5.5) | 4.8<br>(1.5, 8.1) |  |
| Bisexual | 11.2<br>(6.9, 15.4) | 11.0<br>(7.8, 14.3) | 15.1<br>(11.5, 18.7) | 13.0<br>(10.3, 15.6) | 17.3<br>(12.5, 22.2) |  |
| Don't know | 0.34<br>(0.00, 1.01) | 0.38<br>(0.00, 0.96) | 1.3<br>(0.00, 3.0) | 0.33<br>(0.00, 0.90) | 1.3<br>(0.08, 2.6) |  |

|  | 2015 | 2016 | 2017 | 2018 | 2019 | 2015 vs 2019 |
| --- | --- | --- | --- | --- | --- | --- |
| Demographic Characteristics | (N=57,146) | (N=56,897) | (N=56,276) | (N=56,313) | (N=56,136) | *p*-value |
| Race |  |  |  |  |  | **0.04** |
| No LSD Use |  |  |  |  |  |  |
| Non-Hispanic White | 63.7<br>(62.7, 64.6) | 63.3<br>(62.4, 64.2) | 62.7<br>(61.8, 63.6) | 62.3<br>(61.4, 63.2) | 61.9<br>(60.9, 62.9) |  |
| Non-Hispanic Black/African American | 12.0<br>(11.4, 12.6) | 12.0<br>(11.4, 12.6) | 12.1<br>(11.4, 12.8) | 12.1<br>(11.5, 12.7) | 12.2<br>(11.4, 12.9) |  |
| Non-Hispanic Asian | 5.5<br>(5.0, 5.9) | 5.4<br>(5.0, 5.8) | 5.6<br>(5.3, 6.0) | 5.7<br>(5.2, 6.1) | 5.8<br>(5.3, 6.3) |  |
| Hispanic | 16.3<br>(15.6, 16.9) | 16.5<br>(15.9, 17.1) | 16.8<br>(16.0, 17.6) | 17.1<br>(16.3, 17.9) | 17.2<br>(16.5, 18.0) |  |
| Other | 2.6<br>(2.4, 2.8) | 2.8<br>(2.6, 3.0) | 2.7<br>(2.5, 3.0) | 2.9<br>(2.6, 3.1) | 2.9<br>(2.7, 3.1) |  |
| Past-year LSD Use |  |  |  |  |  |  |
| Non-Hispanic White | 71.5<br>(67.0, 76.0) | 71.6<br>(66.2, 77.1) | 72.6<br>(68.0, 77.1) | 70.2<br>(64.8, 75.6) | 69.8<br>(65.2, 74.5) |  |
| Non-Hispanic Black/African American | 5.0<br>(2.9, 7.2) | 3.9<br>(1.8, 6.0) | 4.8<br>(1.8, 7.9) | 4.6<br>(3.0, 6.2) | 4.1<br>(2.3, 5.8) |  |
| Non-Hispanic Asian | 3.4<br>(1.6, 5.2) | 4.1<br>(1.7, 6.4) | 3.4<br>(1.2, 5.6) | 6.1<br>(3.7, 8.4) | 3.8<br>(1.8, 5.8) |  |
| Hispanic | 15.1<br>(11.2, 19.1) | 16.7<br>(12.1, 21.3) | 14.9<br>(11.6, 18.1) | 13.5<br>(10.3, 16.7) | 17.5<br>(13.6, 21.5) |  |
| Other | 4.9<br>(3.0, 6.9) | 3.7<br>(2.3, 5.2) | 4.3<br>(2.2, 6.3) | 5.6<br>(3.1, 8.1) | 4.8<br>(2.3, 7.2) |  |
| Marital status* |  |  |  |  |  | **0.004** |
| No LSD Use |  |  |  |  |  |  |

|  | **2015** | **2016** | **2017** | **2018** | **2019** | **2015 vs 2019** |
| --- | --- | --- | --- | --- | --- | --- |
| **Demographic Characteristics** | **(N=57,146)** | **(N=56,897)** | **(N=56,276)** | **(N=56,313)** | **(N=56,136)** | **p-value** |
| Married | 50.4<br>(49.6, 51.1) | 49.4<br>(48.6, 50.2) | 49.6<br>(48.7, 50.6) | 49.4<br>(48.5, 50.2) | 48.8<br>(48.0, 49.6) |  |
| Widowed | 6.0<br>(5.6, 6.4) | 5.6<br>(5.2, 6.1) | 5.3<br>(4.9, 5.6) | 5.5<br>(5.1, 5.8) | 6.0<br>(5.7, 6.3) |  |
| Divorced or Separated | 13.3<br>(12.8, 13.8) | 13.3<br>(12.8, 13.8) | 13.1<br>(12.6, 13.7) | 13.2<br>(12.7, 13.7) | 13.2<br>(12.6, 13.8) |  |
| Never Been Married | 30.3<br>(29.8, 30.9) | 31.6<br>(30.9, 32.4) | 32.0<br>(31.2, 32.7) | 31.9<br>(31.2, 32.6) | 32.0<br>(31.4, 32.6) |  |
| Past-year LSD Use |  |  |  |  |  |  |
| Married | 5.7<br>(3.0, 8.5) | 5.6<br>(2.4, 8.8) | 8.2<br>(5.0, 11.3) | 7.9<br>(4.2, 11.6) | 10.1<br>(5.0, 15.1) |  |
| Widowed | 1.03<br>(0.05, 2.0) | 0.16<br>(0.00, 0.47) | 1.2<br>(0.00, 3.6) | 0 | 0.29<br>(0.00, 0.87) |  |
| Divorced or Separated | 5.3<br>(1.6, 8.9) | 3.3<br>(1.5, 5.0) | 4.5<br>(1.08, 7.9) | 4.0<br>(1.6, 6.4) | 5.8<br>(3.1, 8.5) |  |
| Never Been Married | 88.0<br>(83.8, 92.1) | 91.0<br>(87.3, 94.6) | 86.1<br>(81.4, 90.8) | 88.1<br>(84.0, 92.2) | 83.9<br>(79.2, 88.5) |  |
| Urbanicity<sup>1</sup> |  |  |  |  |  | **0.94** |
| No LSD Use |  |  |  |  |  |  |
| Large Metro | 55.9<br>(54.8, 57.0) | 55.8<br>(54.9, 56.6) | 56.2<br>(55.5, 56.9) | 55.7<br>(54.7, 56.7) | 55.9<br>(54.9, 56.8) |  |
| Small Metro | 30.4<br>(29.4, 31.5) | 29.9<br>(28.9, 30.8) | 29.2<br>(28.5, 30.0) | 29.9<br>(28.8, 30.9) | 30.3<br>(29.4, 31.2) |  |
| Nonmetro | 13.7<br>(13.0, 14.4) | 14.3<br>(13.5, 15.2) | 14.5<br>(13.8, 15.3) | 14.4<br>(13.6, 15.3) | 13.8<br>(13.0, 14.7) |  |
| Past-year LSD Use |  |  |  |  |  |  |

|  | 2015 | 2016 | 2017 | 2018 | 2019 | 2015 vs 2019 |
| --- | --- | --- | --- | --- | --- | --- |
| Demographic Characteristics | (N=57,146) | (N=56,897) | (N=56,276) | (N=56,313) | (N=56,136) | *p*-value |
| Large Metro | 60.4<br>(54.9, 66.0) | 60.9<br>(55.2, 66.6) | 60.9<br>(55.7, 66.2) | 62.3<br>(57.4, 67.2) | 53.2<br>(48.1, 58.3) |  |
| Small Metro | 29.2<br>(24.7, 33.7) | 28.9<br>(23.1, 34.7) | 27.4<br>(22.6, 32.2) | 28.5<br>(24.2, 32.7) | 32.3<br>(27.7, 36.9) |  |
| Nonmetro | 10.3<br>(6.9, 13.8) | 10.2<br>(7.3, 13.1) | 11.6<br>(8.8, 14.4) | 9.2<br>(6.5, 12.0) | 14.5<br>(10.3, 18.7) |  |
| *p*-value | 0.11c | 0.059c | 0.12c | 0.002c | 0.61c |  |
| Education* |  |  |  |  |  | <.0001 |
| No LSD Use |  |  |  |  |  |  |
| Some high school, no diploma | 22.0<br>(21.5, 22.5) | 20.8<br>(20.3, 21.3) | 20.2<br>(19.5, 20.9) | 20.3<br>(19.8, 20.8) | 19.9<br>(19.3, 20.4) |  |
| High school diploma/GED | 23.1<br>(22.4, 23.9) | 23.0<br>(22.5, 23.6) | 22.2<br>(21.6, 22.8) | 22.7<br>(22.1, 23.3) | 22.2<br>(21.7, 22.8) |  |
| Some college, no de | 19.2<br>(18.6, 19.7) | 19.6<br>(19.1, 20.0) | 19.7<br>(19.1, 20.2) | 19.7<br>(19.2, 20.2) | 19.4<br>(18.8, 19.9) |  |
| 2-year college degr | 8.5<br>(8.1, 8.8) | 8.4<br>(7.9, 8.8) | 8.5<br>(8.1, 8.9) | 8.4<br>(8.0, 8.7) | 8.5<br>(8.1, 8.9) |  |
| 4-year college degr | 27.2<br>(26.4, 28.0) | 28.2<br>(27.4, 29.0) | 29.4<br>(28.4, 30.3) | 28.9<br>(28.0, 29.9) | 30.1<br>(29.3, 30.8) |  |
| Past-year LSD Use |  |  |  |  |  |  |
| Some high school, no diploma | 24.5<br>(20.2, 28.9) | 19.9<br>(16.0, 23.9) | 18.5<br>(15.4, 21.5) | 14.0<br>(10.9, 17.0) | 17.4<br>(14.0, 20.9) |  |
| High school diploma/GED | 21.7<br>(18.0, 25.4) | 22.4<br>(16.7, 28.1) | 22.5<br>(17.7, 27.3) | 17.6<br>(13.9, 21.2) | 22.6<br>(18.0, 27.1) |  |
| Some college, no de | 33.2<br>(27.9, 38.5) | 37.0<br>(31.0, 43.1) | 32.6<br>(27.3, 37.9) | 33.1<br>(28.6, 37.7) | 29.2<br>(24.5, 33.8) |  |

|  | **2015** | **2016** | **2017** | **2018** | **2019** | **2015 vs 2019** |
| --- | --- | --- | --- | --- | --- | --- |
| **Demographic Characteristics** | **(N=57,146)** | **(N=56,897)** | **(N=56,276)** | **(N=56,313)** | **(N=56,136)** | **p-value** |
| 2-year college degr | 5.4<br>(3.0,<br>7.8) | 7.0<br>(4.1,<br>9.8) | 5.3<br>(3.4,<br>7.2) | 6.9<br>(4.2,<br>9.5) | 7.9<br>(5.4,<br>10.4) |  |
| 4-year college degr | 15.1<br>(11.4,<br>18.8) | 13.6<br>(9.5,<br>17.8) | 21.2<br>(16.1,<br>26.3) | 28.5<br>(23.3,<br>33.7) | 22.9<br>(18.2,<br>27.7) |  |
| **Employment status\*** |  |  |  |  |  | **0.003** |
| No LSD Use |  |  |  |  |  |  |
| Employed full time | 46.6<br>(45.9,<br>47.3) | 47.0<br>(46.3,<br>47.7) | 47.6<br>(46.9,<br>48.3) | 47.5<br>(46.7,<br>48.3) | 47.5<br>(46.7,<br>48.3) |  |
| Employed part time | 13.8<br>(13.4,<br>14.2) | 13.5<br>(12.9,<br>14.1) | 13.5<br>(13.1,<br>13.9) | 13.4<br>(13.0,<br>13.8) | 13.4<br>(13.0,<br>13.9) |  |
| Unemployed | 5.0<br>(4.7,<br>5.2) | 4.8<br>(4.5,<br>5.1) | 4.6<br>(4.4,<br>4.9) | 4.3<br>(4.0,<br>4.6) | 4.2<br>(4.0,<br>4.5) |  |
| Other | 34.6<br>(33.8,<br>35.5) | 34.7<br>(34.0,<br>35.3) | 34.3<br>(33.5,<br>35.1) | 34.7<br>(33.9,<br>35.5) | 34.8<br>(34.0,<br>35.6) |  |
| Past-year LSD Use |  |  |  |  |  |  |
| Employed full time | 44.5<br>(39.2,<br>49.8) | 42.6<br>(36.5,<br>48.6) | 47.9<br>(42.3,<br>53.4) | 50.3<br>(46.1,<br>54.6) | 45.0<br>(39.5,<br>50.4) |  |
| Employed part time | 27.4<br>(22.1,<br>32.7) | 29.8<br>(24.2,<br>35.3) | 22.5<br>(17.8,<br>27.2) | 25.8<br>(21.3,<br>30.2) | 26.4<br>(22.0,<br>30.8) |  |
| Unemployed | 8.0<br>(5.4,<br>10.6) | 11.8<br>(8.3,<br>15.4) | 11.5<br>(7.7,<br>15.4) | 9.6<br>(6.4,<br>12.7) | 10.7<br>(7.6,<br>13.7) |  |
| Other | 20.1<br>(16.0,<br>24.1) | 15.8<br>(12.1,<br>19.6) | 18.1<br>(14.6,<br>21.6) | 14.4<br>(11.4,<br>17.3) | 18.0<br>(14.0,<br>22.0) |  |
| **Past 12 mos, how many employers, Age 18+ only** |  |  |  |  |  | **0.30** |
| No LSD Use |  |  |  |  |  |  |

|  | **2015** | **2016** | **2017** | **2018** | **2019** | **2015 vs 2019** |
| --- | --- | --- | --- | --- | --- | --- |
| **Demographic Characteristics** | **(N=57,146)** | **(N=56,897)** | **(N=56,276)** | **(N=56,313)** | **(N=56,136)** | **p-value** |
| One | 75.4<br>(74.7, 76.1) | 74.3<br>(73.6, 75.0) | 74.9<br>(74.3, 75.6) | 75.0<br>(74.3, 75.6) | 74.8<br>(74.1, 75.6) |  |
| Two | 18.1<br>(17.6, 18.7) | 18.5<br>(17.9, 19.1) | 17.7<br>(17.2, 18.3) | 18.4<br>(17.7, 19.0) | 18.3<br>(17.7, 18.9) |  |
| Three | 4.2<br>(4.0, 4.4) | 4.5<br>(4.3, 4.8) | 4.5<br>(4.2, 4.8) | 4.2<br>(3.9, 4.5) | 4.4<br>(4.1, 4.7) |  |
| Four or more | 2.3<br>(2.1, 2.5) | 2.6<br>(2.4, 2.8) | 2.9<br>(2.6, 3.1) | 2.4<br>(2.2, 2.6) | 2.5<br>(2.2, 2.7) |  |
| Past-year LSD Use |  |  |  |  |  |  |
| One | 51.7<br>(45.8, 57.6) | 43.0<br>(36.7, 49.4) | 45.6<br>(39.7, 51.4) | 49.7<br>(43.6, 55.9) | 54.1<br>(47.9, 60.3) |  |
| Two | 31.1<br>(26.3, 36.0) | 36.8<br>(29.4, 44.2) | 34.0<br>(28.6, 39.5) | 29.8<br>(24.3, 35.3) | 26.7<br>(22.0, 31.4) |  |
| Three | 11.8<br>(7.5, 16.1) | 13.8<br>(10.0, 17.5) | 13.8<br>(10.2, 17.3) | 13.7<br>(9.9, 17.4) | 11.9<br>(8.7, 15.1) |  |
| Four or more | 5.4<br>(3.0, 7.8) | 6.4<br>(4.0, 8.8) | 6.6<br>(4.1, 9.2) | 6.8<br>(4.4, 9.2) | 7.3<br>(4.4, 10.3) |  |
| **Total family income\*** |  |  |  |  |  | **<.0001** |
| No LSD Use |  |  |  |  |  |  |
| Less than \$20,000 | 17.7<br>(17.1, 18.3) | 16.8<br>(16.1, 17.4) | 16.0<br>(15.3, 16.7) | 15.6<br>(15.0, 16.2) | 14.6<br>(14.1, 15.1) | |
| \$20,000 - \$49,999 | 29.8<br>(29.1, 30.6) | 29.8<br>(29.1, 30.6) | 29.3<br>(28.5, 30.0) | 29.2<br>(28.5, 29.8) | 28.3<br>(27.4, 29.1) | |
| \$50,000 - \$74,999 | 16.5<br>(15.9, 17.0) | 15.8<br>(15.2, 16.4) | 15.7<br>(15.2, 16.2) | 15.4<br>(15.0, 15.9) | 15.8<br>(15.3, 16.3) | |

|  | **2015** | **2016** | **2017** | **2018** | **2019** | **2015 vs 2019** |
| --- | --- | --- | --- | --- | --- | --- |
| **Demographic Characteristics** | **(N=57,146)** | **(N=56,897)** | **(N=56,276)** | **(N=56,313)** | **(N=56,136)** | ***p*-value** |
| \$75,000 or More | 36.0<br>(35.1, 36.9) | 37.6<br>(36.6, 38.5) | 39.0<br>(38.1, 40.0) | 39.8<br>(38.9, 40.8) | 41.3<br>(40.3, 42.3) | |
| Past-year LSD Use |  |  |  |  |  |  |
| Less than \$20,000 | 23.9<br>(19.2, 28.6) | 27.5<br>(22.3, 32.6) | 28.8<br>(23.6, 33.9) | 24.4<br>(19.6, 29.1) | 22.8<br>(18.3, 27.2) | |
| \$20,000 - \$49,999 | 33.3<br>(28.7, 38.0) | 27.7<br>(23.5, 32.0) | 24.8<br>(21.1, 28.4) | 32.7<br>(28.5, 36.8) | 30.4<br>(25.9, 34.8) | |
| \$50,000 - \$74,999 | 12.9<br>(8.6, 17.1) | 17.3<br>(13.1, 21.4) | 16.5<br>(12.8, 20.2) | 11.6<br>(8.2, 15.0) | 14.9<br>(11.5, 18.3) | |
| \$75,000 or More | 29.9<br>(24.7, 35.1) | 27.6<br>(22.1, 33.0) | 30.0<br>(25.3, 34.7) | 31.4<br>(26.4, 36.3) | 32.0<br>(26.5, 37.5) | |
| Poverty level (% of US census poverty threshold)* |  |  |  |  |  | **<.0001** |
| No LSD Use |  |  |  |  |  |  |
| Living in Poverty | 15.9<br>(15.4, 16.4) | 15.1<br>(14.6, 15.7) | 14.5<br>(13.9, 15.1) | 14.6<br>(14.1, 15.1) | 13.6<br>(13.2, 14.1) |  |
| Income ↑ 2X Fed Pov Thresh | 20.4<br>(19.7, 21.1) | 20.4<br>(19.8, 21.0) | 20.1<br>(19.4, 20.7) | 19.9<br>(19.3, 20.4) | 19.2<br>(18.6, 19.8) |  |
| Income > 2X Fed Pov Thresh | 63.7<br>(62.9, 64.6) | 64.5<br>(63.7, 65.3) | 65.5<br>(64.5, 66.5) | 65.6<br>(64.7, 66.4) | 67.2<br>(66.3, 68.1) |  |
| Past-year LSD Use |  |  |  |  |  |  |
| Living in Poverty | 18.4<br>(14.6, 22.1) | 21.3<br>(17.3, 25.4) | 23.5<br>(19.0, 28.0) | 19.7<br>(15.8, 23.6) | 19.2<br>(15.2, 23.1) |  |
| Income ↑ 2X Fed Pov Thresh | 22.4<br>(17.7, 27.1) | 22.2<br>(17.0, 27.3) | 19.4<br>(15.7, 23.1) | 23.8<br>(19.3, 28.2) | 22.1<br>(18.1, 26.1) |  |
| Income > 2X Fed Pov Thresh | 59.2<br>(54.0, 64.4) | 56.5<br>(51.9, 61.1) | 57.1<br>(51.3, 62.9) | 56.5<br>(50.4, 62.7) | 58.7<br>(53.4, 64.1) |  |

|  | **2015** | **2016** | **2017** | **2018** | **2019** | **2015 vs 2019** |
| --- | --- | --- | --- | --- | --- | --- |
| **Demographic Characteristics** | **(N=57,146)** | **(N=56,897)** | **(N=56,276)** | **(N=56,313)** | **(N=56,136)** | **p-value** |
| Are you pregnant<sup>2</sup> |  |  |  |  |  | **0.42** |
| No LSD Use | 3.3<br>(3.0, 3.6) | 3.3<br>(3.0, 3.6) | 3.3<br>(3.0, 3.6) | 3.5<br>(3.2, 3.8) | 2.9<br>(2.6, 3.2) |  |
| Past-year LSD Use | 1.1<br>(0.00, 2.6) | 0.80<br>(0.00, 1.9) | 1.6<br>(0.00, 3.6) | 3.0<br>(0.00, 6.1) | 2.1<br>(0.00, 4.3) |  |
| # Religious services past 12 months<sup>3</sup> |  |  |  |  |  | **0.01** |
| No LSD Use |  |  |  |  |  |  |
| 25 or More | 67.3<br>(66.7, 67.9) | 67.5<br>(66.9, 68.0) | 68.3<br>(67.5, 69.1) | 68.7<br>(67.9, 69.4) | 68.5<br>(67.8, 69.2) |  |
| Less than 25 | 32.7<br>(32.1, 33.3) | 32.5<br>(32.0, 33.1) | 31.7<br>(30.9, 32.5) | 31.3<br>(30.6, 32.1) | 31.5<br>(30.8, 32.2) |  |
| Past-year LSD Use |  |  |  |  |  |  |
| 25 or More | 66.3<br>(61.4, 71.2) | 69.5<br>(64.7, 74.2) | 68.8<br>(64.7, 72.9) | 71.2<br>(66.4, 76.0) | 71.9<br>(67.1, 76.8) |  |
| Less than 25 | 33.7<br>(28.8, 38.6) | 30.5<br>(25.8, 35.3) | 31.2<br>(27.1, 35.3) | 28.8<br>(24.0, 33.6) | 28.1<br>(23.2, 32.9) |  |
| My religious beliefs influence my decisions |  |  |  |  |  | **<.0001** |
| No LSD Use |  |  |  |  |  |  |
| Strongly Disagree | 16.0<br>(15.5, 16.6) | 16.9<br>(16.4, 17.4) | 17.3<br>(16.7, 17.9) | 17.7<br>(17.4, 18.1) | 18.3<br>(17.7, 18.9) |  |
| Disagree | 15.2<br>(14.6, 15.7) | 15.6<br>(15.2, 16.1) | 15.6<br>(15.1, 16.1) | 15.7<br>(15.2, 16.1) | 16.2<br>(15.8, 16.6) |  |
| Agree | 35.8<br>(35.1, 36.5) | 34.9<br>(34.3, 35.6) | 35.1<br>(34.6, 35.7) | 35.6<br>(34.9, 36.2) | 34.7<br>(34.0, 35.4) |  |
| Strongly Agree | 33.0<br>(32.3, 33.6) | 32.5<br>(31.9, 33.2) | 32.0<br>(31.4, 32.6) | 31.0<br>(30.4, 31.6) | 30.8<br>(30.1, 31.5) |  |

|  | **2015** | **2016** | **2017** | **2018** | **2019** | **2015 vs 2019** |
| --- | --- | --- | --- | --- | --- | --- |
| **Demographic Characteristics** | **(N=57,146)** | **(N=56,897)** | **(N=56,276)** | **(N=56,313)** | **(N=56,136)** | **p-value** |
| Past-year LSD Use |  |  |  |  |  |  |
| Strongly Disagree | 45.5<br>(39.5, 51.5) | 46.6<br>(40.3, 52.9) | 43.2<br>(38.1, 48.3) | 45.6<br>(39.7, 51.5) | 47.6<br>(41.4, 53.7) |  |
| Disagree | 26.8<br>(21.5, 32.1) | 21.8<br>(17.5, 26.1) | 23.2<br>(19.8, 26.7) | 23.4<br>(20.0, 26.7) | 23.3<br>(18.8, 27.9) |  |
| Agree | 20.8<br>(16.9, 24.8) | 23.9<br>(18.3, 29.4) | 26.5<br>(21.8, 31.2) | 24.6<br>(18.9, 30.4) | 20.7<br>(16.6, 24.8) |  |
| Strongly Agree | 6.9<br>(3.3, 10.4) | 7.8<br>(4.6, 10.9) | 7.1<br>(4.8, 9.3) | 6.5<br>(3.8, 9.2) | 8.4<br>(5.5, 11.4) |  |
| My religious beliefs are very important |  |  |  |  |  | **<.0001** |
| No LSD Use |  |  |  |  |  |  |
| Strongly Disagree | 16.3<br>(15.8, 16.9) | 16.9<br>(16.4, 17.4) | 17.6<br>(17.0, 18.1) | 18.2<br>(17.7, 18.6) | 18.3<br>(17.8, 18.9) |  |
| Disagree | 12.3<br>(11.9, 12.7) | 13.0<br>(12.6, 13.5) | 12.8<br>(12.4, 13.3) | 12.8<br>(12.3, 13.2) | 13.4<br>(13.0, 13.9) |  |
| Agree | 34.1<br>(33.4, 34.7) | 33.2<br>(32.7, 33.8) | 33.3<br>(32.6, 33.9) | 33.8<br>(33.2, 34.5) | 33.7<br>(33.1, 34.3) |  |
| Strongly Agree | 37.3<br>(36.5, 38.0) | 36.8<br>(36.1, 37.5) | 36.4<br>(35.7, 37.0) | 35.2<br>(34.6, 35.8) | 34.5<br>(33.9, 35.1) |  |
| Past-year LSD Use |  |  |  |  |  |  |
| Strongly Disagree | 43.5<br>(38.2, 48.9) | 44.1<br>(37.0, 51.2) | 42.2<br>(37.4, 47.0) | 43.5<br>(37.6, 49.5) | 45.2<br>(39.2, 51.3) |  |
| Disagree | 23.2<br>(18.1, 28.2) | 22.6<br>(17.5, 27.6) | 22.0<br>(17.5, 26.5) | 22.1<br>(18.5, 25.6) | 21.8<br>(18.1, 25.6) |  |
| Agree | 25.4<br>(21.2, 29.6) | 21.3<br>(16.1, 26.4) | 25.8<br>(20.7, 30.8) | 25.8<br>(21.4, 30.2) | 21.3<br>(16.6, 25.9) |  |

|  | 2015 | 2016 | 2017 | 2018 | 2019 | 2015 vs 2019 |
| --- | --- | --- | --- | --- | --- | --- |
| Demographic Characteristics | (N=57,146) | (N=56,897) | (N=56,276) | (N=56,313) | (N=56,136) | *p*-value |
| Strongly Agree | 7.9<br>(4.4, 11.3) | 12.1<br>(7.6, 16.5) | 10.0<br>(7.3, 12.7) | 8.6<br>(5.3, 11.8) | 11.7<br>(7.7, 15.7) |  |
| # Days skipped work past 30 days, median [Q1, Q3] |  |  |  |  |  |  |
| No LSD Use | 0.00<br>[0.00, 0.00] | 0.00<br>[0.00, 0.00] | 0.00<br>[0.00, 0.00] | 0.00<br>[0.00, 0.00] | 0.00<br>[0.00, 0.00] | <.0001 |
| Past-year LSD Use | 0.00<br>[0.00, 0.26] | 0.00<br>[0.00, 0.00] | 0.00<br>[0.00, 0.31] | 0.00<br>[0.00, 0.51] | 0.00<br>[0.00, 0.05] |  |
| Body mass index (BMI) (Age 18+ only), median [Q1, Q3]* |  |  |  |  |  | <.0001 |
| No LSD Use | 26.6<br>[23.2, 31.2] | 26.7<br>[23.3, 31.2] | 26.6<br>[23.2, 31.3] | 26.9<br>[23.3, 31.4] | 27.0<br>[23.4, 31.6] |  |
| Past-year LSD Use | 23.8<br>[21.3, 27.0] | 23.9<br>[21.6, 27.7] | 23.5<br>[20.9, 27.4] | 23.4<br>[21.0, 26.9] | 23.7<br>[21.0, 27.0] |  |

Frequencies presented are unweighted counts. Weighted proportions and 95% confidence intervals are reported.
\*Imputed or recoded variable.
Reported P-values: a=linear regression; b=linear regression with log transformation; c=Rao-Scott chi-square test.
<sup>1</sup>The association between Past-Year LSD use and Urbanicity was only significant in 2018.
<sup>2</sup>The association between Past-Year LSD use and Pregnancy was only significant in 2016.
<sup>3</sup>There was no association between Past-Year LSD use and the number of religious services attended within any survey year. All other covariates were significantly associated with past-year LSD use within each survey year ( $p < 0.001$ ).
The associations remained stable over time for all covariates except age, marital status, and pregnancy (all $p < 0.001$ for the interaction term with year).
All associations are adjusted for survey year, age, employment, education, race, metropolitan area, and religious importance. SAS Survey Procedures used for all analyses.

Associations between past-year LSD use with the use of other drugs were consistent across the study period, except for rising past-year LSD use among lifetime users of methamphetamine (22.6% in 2019 vs 14.0% in 2014, p=0.04). There was no statistically significant change in the proportion of people with hallucinogen use disorder reporting past-year LSD use (p=0.42). For more information see Supplemental table 3.

### Multivariable modelling of risk for past-year LSD use

In the all ages past-year LSD use multivariable model, factors associated with higher risk of past-year LSD use were: more recent survey year (p<0.001), age 12-17 (p=0.007), part time employment (p<0.001), non-Hispanic Asian race (p=0.04), non-influential and unimportant religious beliefs (p<0.001), having sold illegal drugs at least once (p<0.001), lifetime use of alcohol, marijuana, stimulant, sedatives or any hallucinogen assessed besides LSD (p<0.001), having been approached by someone selling illegal drugs in the past month (p<0.001), lower perceived risk of trying LSD once or twice (p<0.001), and reported easier access to LSD (p<0.001).

In the under age 18 multivariable model, factors associated with higher risk of past-year LSD use are: non-influential and unimportant religious beliefs (p=0.02), overnight stay in a hospital during the past year (p=0.01), lifetime use of tobacco, alcohol, marijuana, stimulant, sedative, or any hallucinogen assessed besides LSD (p<0.001), being approached by someone selling illegal drugs in the past month (p=0.02), lower perceived risk of trying LSD once or twice (p<0.001), and reporting easier access to LSD (p<0.001).

In the over age 18 multivariable model, factors associated with higher risk of past-year LSD use were: more recent survey year (p=0.001 for 2017 and 2018, p<0.001 for 2019), part time employment (p<0.001), non-Hispanic Asian race (p=0.02), non-influential and unimportant religious beliefs (p<0.001), having sold illegal drugs at least once (p<0.001), past-year suicide attempt (p=0.01), lifetime use of marijuana, stimulants, sedatives, or any hallucinogen assessed besides LSD (p<0.001), having been approached by someone selling illegal drugs in the past month (p<0.001), lower perceived risk of trying LSD once or twice (p<0.001), and reported easier access to LSD (p<0.001).

Factors associated with decreased risk of past-year LSD use were similar in all models. In the all-age and over-18 models the factors associated with decreased risk are: marriage (p<0.001), female gender (p<0.001), income more than twice the federal poverty threshold (p<0.001), and having children under 18 years old in the home (p<0.001). In the all-ages model, older respondents (age >18, p<0.001) were less likely to try LSD. In the under-18 model, female gender (p=0.004) was associated with decreased risk of LSD use. For details of these models see Table 3.

**Table 3.**
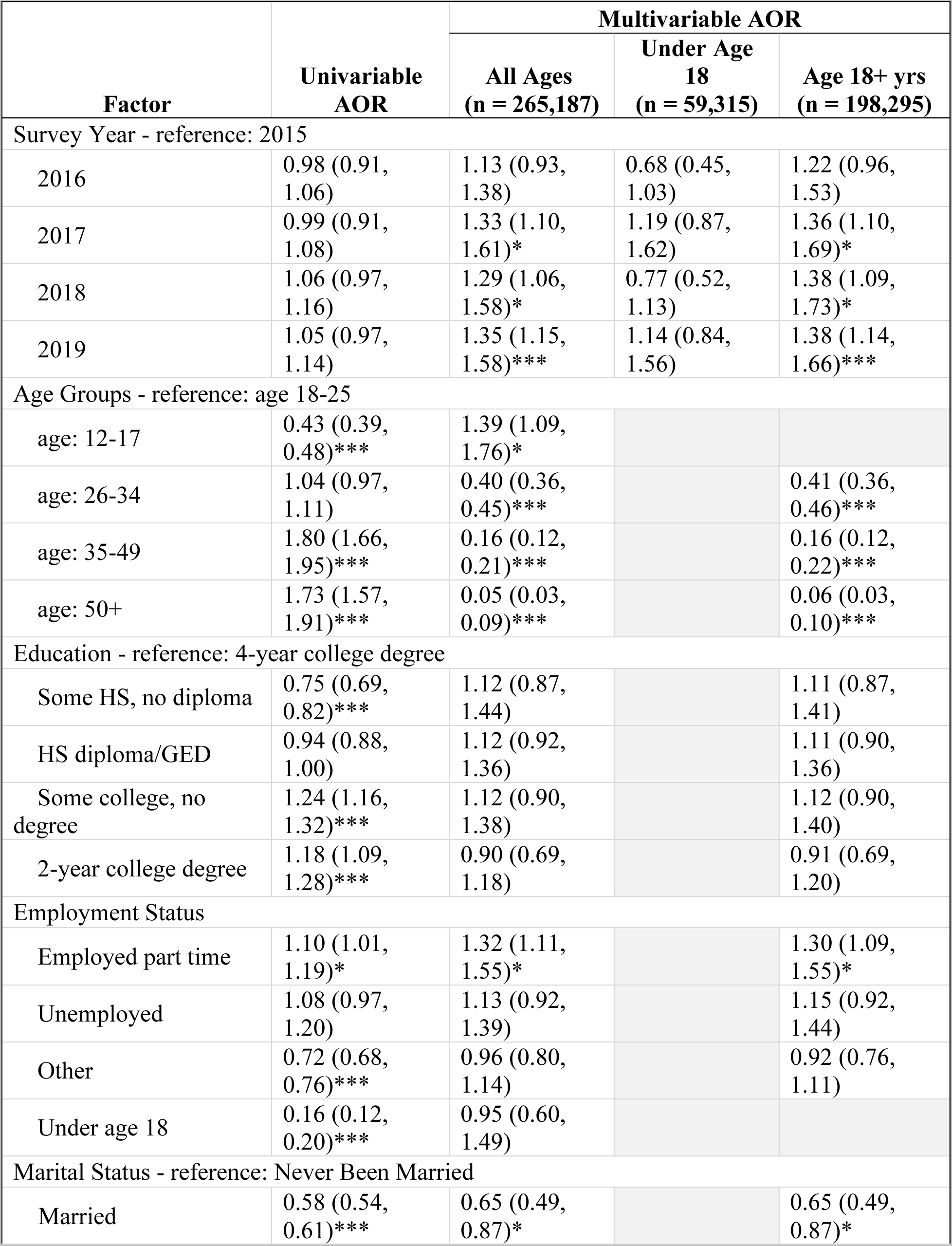
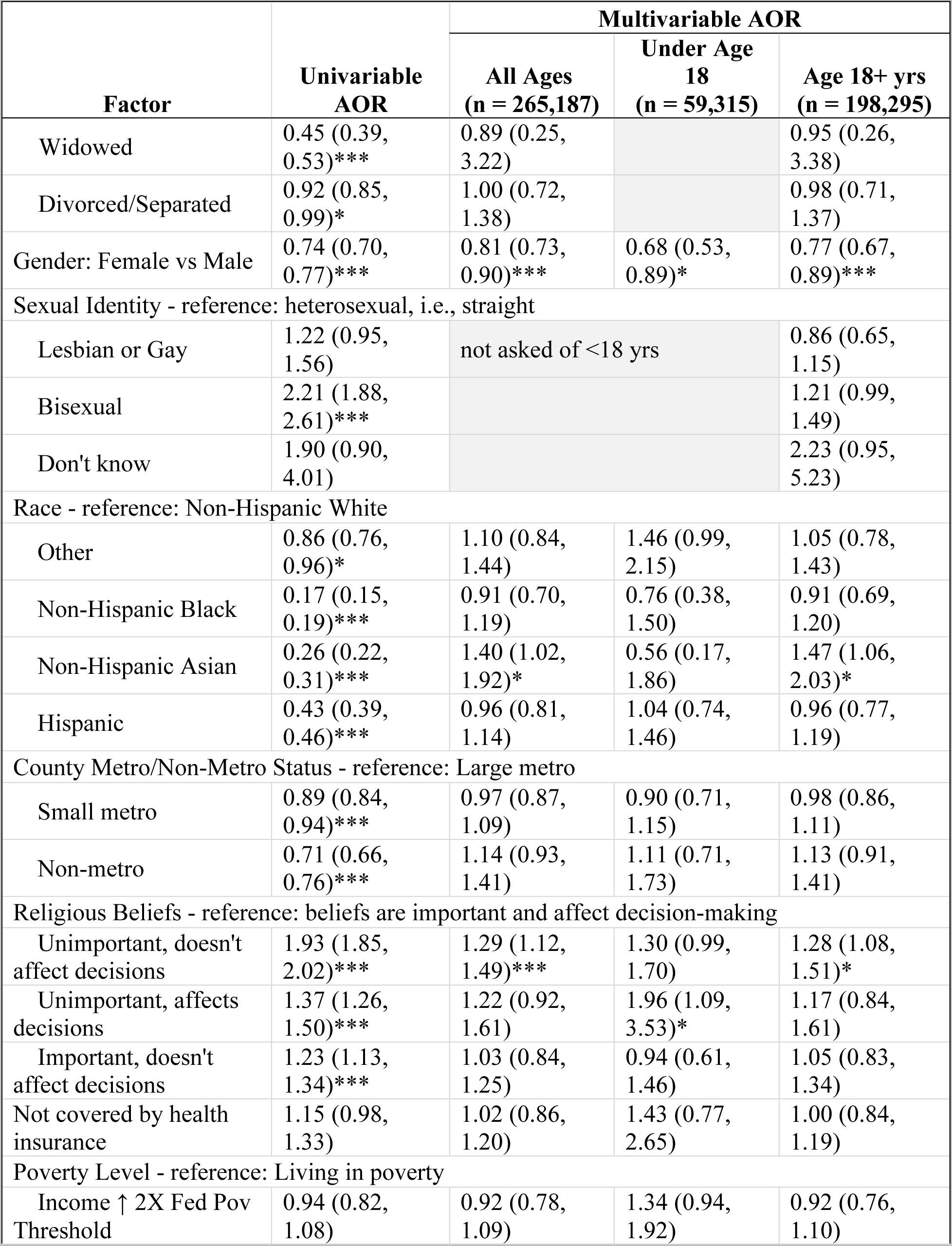
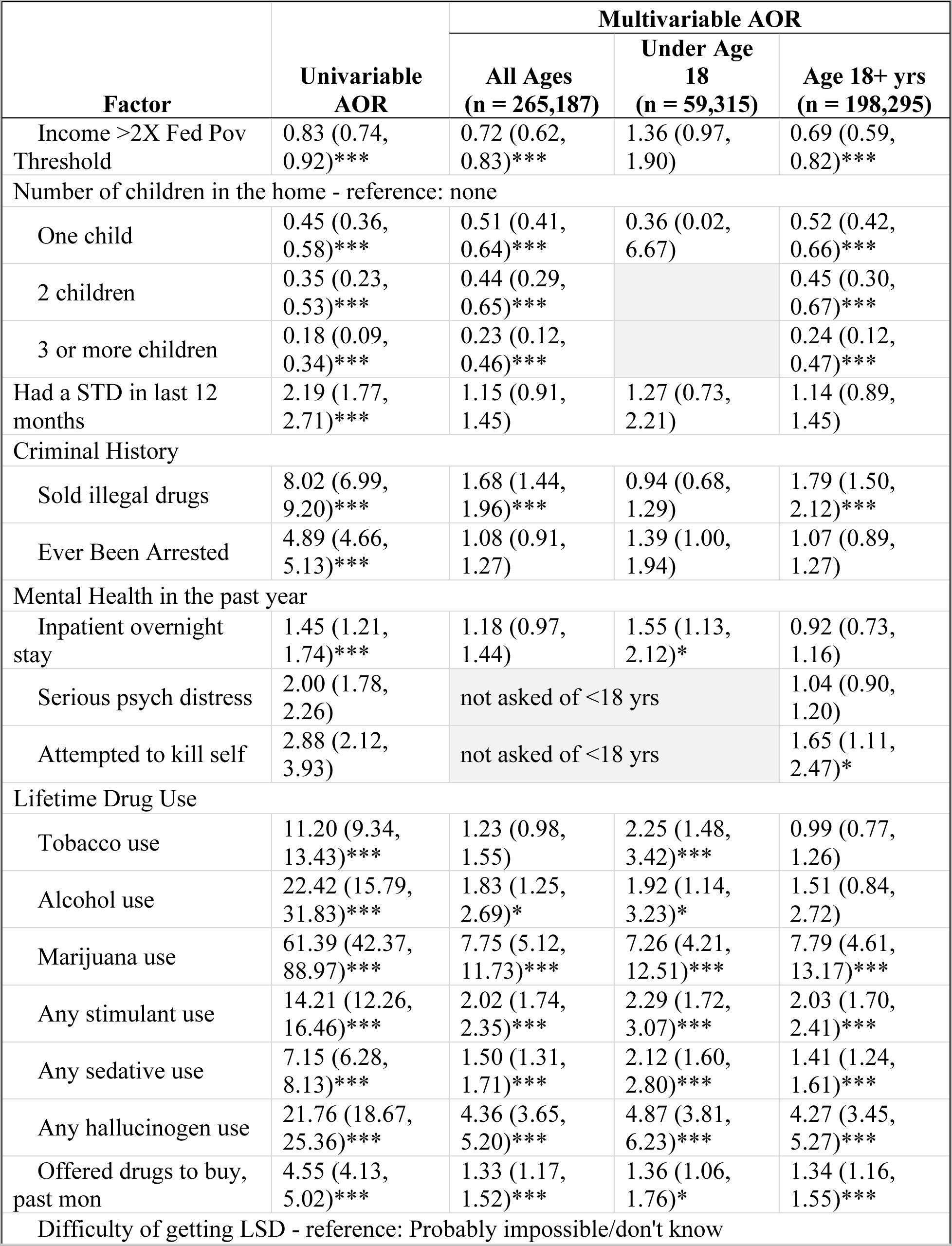
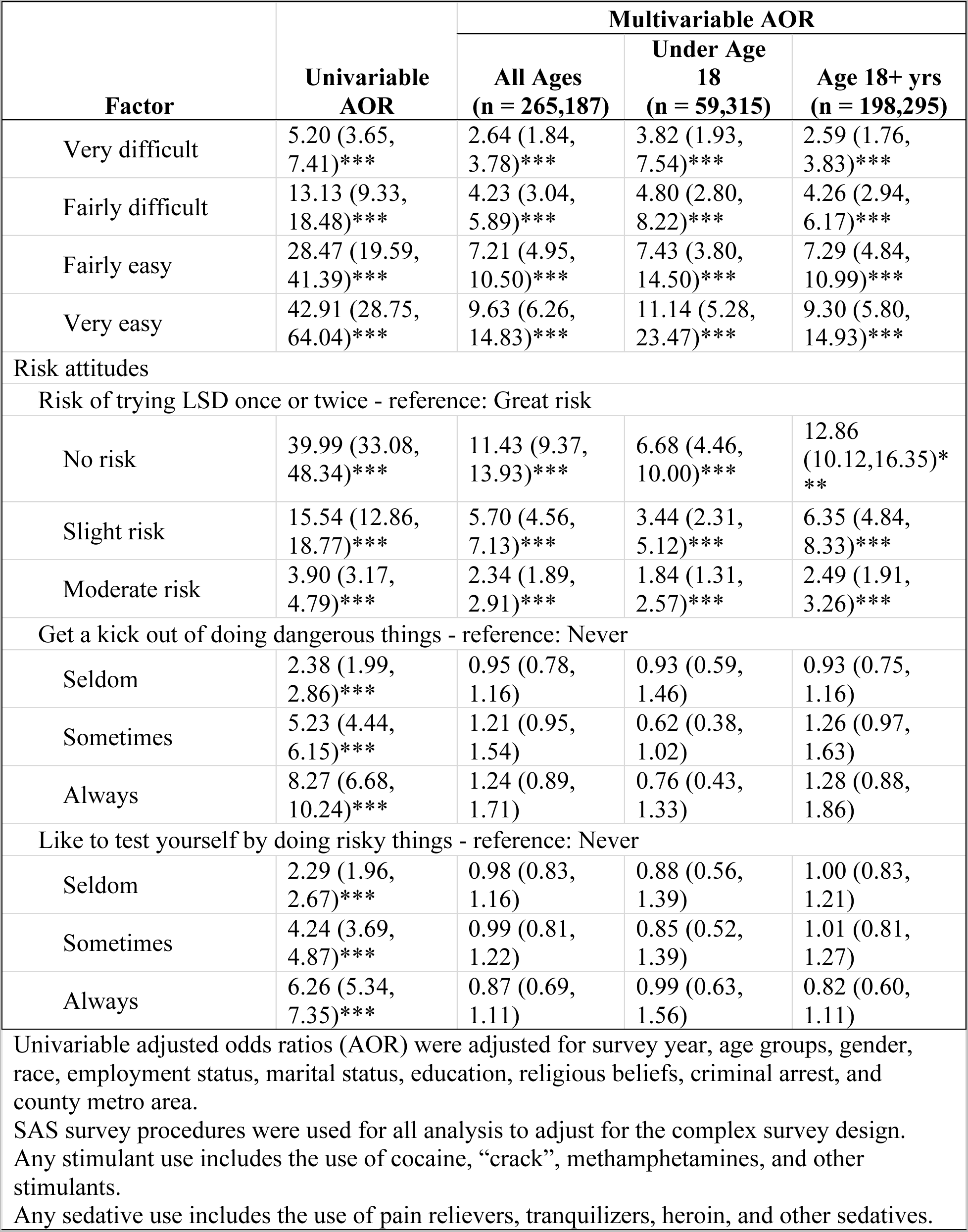
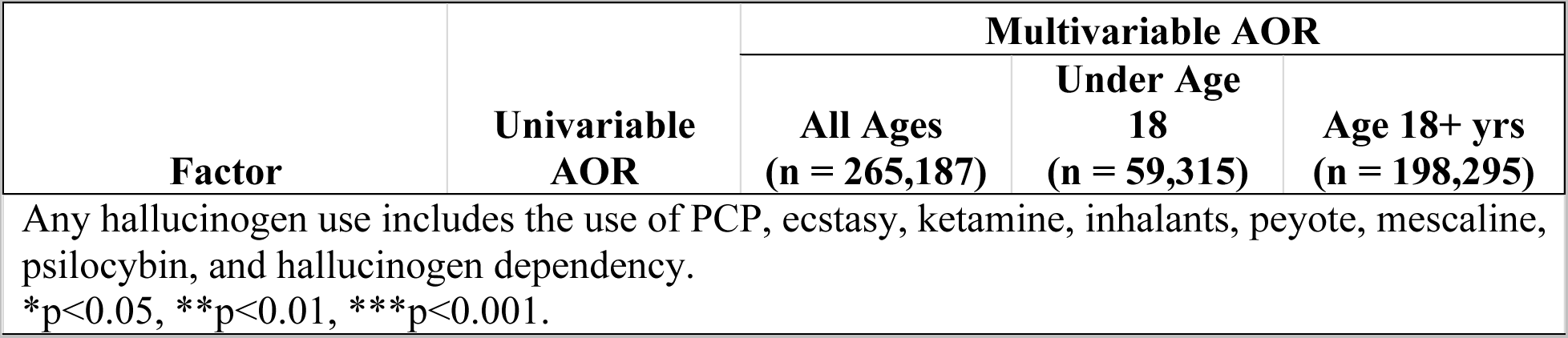
Univariable and Multivariable Adjusted Odds Ratios for the Risk of Past-Year LSD Use from NSDUH Survey Respondents from 2015-2019.

## Discussion

This study provides a detailed look at past-year LSD users in the U.S. from 2015 to 2019. Our findings support other recent findings indicating growing LSD use in the United States (2, 3), with past-year LSD use increasing by 47% from 0.59% to 0.87% between 2015 and 2019. Considering that 2019 past-year use prevalence was 65.1% for alcohol, 26.2% tobacco products, 17.5% for marijuana, 3.7% for opioids, 2.0% cocaine, 1.9% tranquilizers, and 1.8% for stimulants (1, 14), LSD remains an uncommonly used drug in the United States. Notably, despite recent increases in past-year LSD use there was not a statistically significant increase in the proportion of past-year LSD users among people with lifetime history of hallucinogen use disorder, suggesting that other hallucinogens may be more strongly associated with that diagnosis than LSD.

On multivariable analysis, we found that the people most likely to use LSD in the past year were unmarried, non-Hispanic white or Asian men under the age of 26, with no children in the home, with previous alcohol and other substance use, who are employed part time, earning less than twice the federal poverty threshold, consider their religious views to be unimportant, perceive LSD use to be low risk, and have increased access to LSD. Education level appears unrelated to risk of LSD use, as does urbanicity, enjoyment of risky activities, and sexual identity. Past-year LSD use’s lack of association with previous arrest history or past-year sexually transmitted infection (STI), hospitalization, and psychological distress suggests that LSD does not significantly contribute to crime, unsafe sexual practices, or psychiatric problems in the US, though an important exception requiring further exploration was the association between past-year LSD use and past-year suicide attempt in people 18 and older. While we are unaware of reports of suicide attempts in recent clinical trials of LSD (15–18), a large survey of clinicians and researchers treating patients with LSD reported no suicide attempts in research subjects, but 1.2 for every 1,000 patients treated in the community (19). While no conclusion can be made about causality from the data, it is possible that some individuals using LSD may be vulnerable to suicide attempts following use. However, it is also possible that people prone to suicide attempts or who have recently had one are using LSD to self-treat mental illness, perhaps due to recent favorable media coverage of psychedelic-assisted therapy clinical trials. Unfortunately, information about the temporal relationship between past-year LSD use and suicide attempt, as well as motivators for LSD use, are not collected by the NSDUH, preventing clarification on this topic.

We found that people more likely to use LSD have considerable lifetime drug experience, regardless of age. Notably, the proportion of people who had not used LSD in the past year who believed that using LSD once or twice a week was a “great risk” was more than twice as high as the proportion who believed that using marijuana or having five or more alcoholic drinks once or twice a week was a “great risk.” Only a slightly smaller proportion of non-LSD users believed that using LSD once or twice a week was a “great risk” compared to the proportions believing that using cocaine or heroin twice a week was a “great risk.” These findings suggest that LSD is still considered a “hard core” drug by most of the population, despite users only rarely needing emergency medical treatment following use (20). Interestingly, while perceived risk of LSD was predictably lower in users, most users still considered using LSD once or twice a week to be a “moderate” or “great” risk. This might speak to the sometimes-unpredictable nature of psychedelic experience, which may contribute to LSD’s low addictive potential.

Intriguingly, we found that people whose religious views were not important to their lives and decision-making were at increased risk of past-year LSD use. This was surprising given LSD’s ability to induce mystical experiences (21), which can be transformative in some users (22). This suggests that any enhancement of religious or spiritual beliefs occasioned by LSD may be transient in nature. This finding contrasts with that of a small study of people from Israel and Australia that showed on univariate analysis that users of LSD, mescaline, and psilocybin, as a group, more highly valued their spirituality than non-users according to scores on the Life Values Inventory (LVI) (23). There are a variety of possible reasons for this difference, including cultural factors, conceptual differences between religious and spiritual beliefs, and the specific questions asked of participants (the LVI asks participants to rank how strongly “Living in harmony with my spiritual beliefs”, “Believing in a higher power” and “Believing that there is something greater than ourselves” guides their behavior, while the NSDUH asks for level of agreement with the statements “Your religious beliefs are a very important part of your life” and “Your religious beliefs influence how you make decisions in your life”), LSD users being combined with users of other psychedelics in the other study, and the fact that our analysis was multivariable and may have adjusted for confounders. The other study also counted individuals who had used small doses of psychedelics as non-users, whereas the NSDUH does not query about dosage.

While we found a decreased risk of LSD use among Black participants on univariate analysis, we found no statistically significant association between being Black and LSD use on multivariate analysis, suggesting confounding factors may explain this previous finding. Previous research found that Black people may be less likely to use LSD and other psychedelics than White people (24), though this study did not control for confounders. Increased risk for lifetime psilocybin use was observed in bisexual participants of a recent study also employing NSDUH data (25), possibly secondary to efforts by bisexual users to employ psilocybin as a means of coping with the effects of minority stress. In contrast, we identified no relationship between past-year LSD use and sexual identity on multivariable analysis. Regarding another aspect of sexual health, we found no association between past-year LSD use and past-year and STIs. Notably, a 2010 study using NSDUH data found that duration of hallucinogen use was positively associated with risk of lifetime STI diagnosis (26).

The differences in correlates between our under age 18 and our 18+ models included increased odds of LSD use among participants under 18 who reported their religious views were not a very important part of their life but still influenced decision-making, overnight stay in a hospital during the past year, and lifetime tobacco use. Unlike in adults, there was no associations with LSD use among participants under 18 who were non-Hispanic Asian race, had higher income, had children in the home, or had religious beliefs were that were unimportant and did not influence their decision-making.

Finally, we observed a small number of demographic changes in past-year LSD users from 2015-2019. The proportion of LSD users among lifetime users of all substances analyzed remained unchanged during the study period, except for methamphetamine. Methamphetamine use grew during the study period, and its use is associated with polysubstance use, including growing co-occurring opioid use (27, 28) and use among people who use LSD (29). It is unknown whether our finding represents increase simultaneous use of methamphetamine and LSD. Combining LSD with 3,4-Methylenedioxymethamphetamine (MDMA) for synergistic MDMA effects is well known and referred to as “candyflipping” (30, 31). However, combining other stimulants appears to be less common (32). In the case of methamphetamine, this may be due to an increased risk of “bad trips” (33) or diminution of LSD’s effects (34). As observed in another recent study (2), we found that marriage may be a protective factor against LSD use, though it could also be that problematic LSD use or associated drug use produces this finding by contributing to divorce. However, the proportion of married LSD users nearly doubled from 5.7% 2015 to 10.1% in 2019. Though most LSD users remain between the ages of 18 and 25, the proportion of LSD users aged 26-34 grew from 16.3 to 26.5% over the study period. Finally, the proportion of pregnant NSDUH participants reporting past-year LSD use nearly doubled from 1.1% in 2015 to 2.1% in 2019. Though we cannot definitively say this translates into increased use of LSD during pregnancy, this could possibly be the case. Given conflicting evidence on the teratogenic potential of LSD (35), this finding warrants further investigation. Coupled with an overall increase in past-year LSD use during the study period, growth in use among married people, people aged 26-34, and pregnant women, suggests that though LSD remains uncommonly used, societal acceptance in the United States may be growing.

### Strengths and Limitations

The primary strength of this study is its use of data from multiple administrations of a large, rigorously conducted survey employing a nationally representative survey sample. Limitations include the NSDUH’s use of retrospective self-report by participants, which could result in underreporting of LSD and other substance use. However, the NSDUH’s substance use self-report measures have high concordance with drug testing results (36). NSDUH participation restriction to the civilian non-institutionalized population is another important limitation. While this represents 97% of the US population (13), the NSDUH excludes people living in institutional group quarters such as hospitals, prisons, nursing homes, and addiction treatment centers, who are likely to have important differences in past-year LSD use and demographics. Further limitations include the fact that the NSDUH does not report number of LSD exposures, doses of LSD used, setting of use, or reasons for LSD use.

## Conclusions

Past-year LSD use rose 47% from 2015-2019 in the US, though LSD continues to be used by only a sliver of the US population each year. Use is strongly associated with decreased risk perception around LSD and increased ease of access. Non-users of LSD still consider regular use of LSD to be much riskier than regular use of alcohol or marijuana, though slightly less risky than regular use of cocaine or heroin. With increases in the proportion of past-year LSD users aged 26-34, married, and pregnant, we may be seeing the early stages of increased social acceptance of LSD use. We found no associations on multivariable analysis with unemployment, arrest history, past-year psychological distress, or STIs, suggesting that LSD does not significantly contribute significantly to public health problems in the US.

## Data Availability

Publicly available data (NSDUH)

## Acknowledgements

This work was supported by a non-monetary scientific collaboration award from the Cleveland Clinic Center for Populations Health Research awarded to Dr. Barnett.

## Disclosure of relationships and activities

Dr. Barnett has received stock options from CB Therapeutics as compensation for advisory services. He also receives monetary compensation for editorial work for DynaMed Plus (EBSCO Industries, Inc) and consulting services for Cerebral.

## Supplement

### Appendix A: Statistical analysis details

Respondents under the age of 18 were not asked survey questions in areas of mental health (ex. attempted suicide) and certain demographics (ex. sexual identity). Additionally, some survey questions were answered overwhelmingly with a single choice in that age group: married status (99% never been married), education (99% some high school but no diploma), employment status (70% unemployed or other), and number of children present in the household (either 0 or 1 only). Analyses on survey questions restricted by age (over/under age 18) were appropriately stratified.

Regarding variable selection for our analyses, we largely selected variables that had previously been associated with LSD use including: year of survey administration (Killion et al. 2021; R. A. Yockey, Vidourek, and King 2020), age (Killion et al. 2021), education level (Killion et al. 2021), marital status (Killion et al. 2021), gender (Killion et al. 2021; A. Yockey, King, and Vidourek 2019), income (Killion et al. 2021), urbanicity (Killion et al. 2021; A. Yockey, King, and Vidourek 2019), race/ethnicity (Killion et al. 2021; A. Yockey, King, and Vidourek 2019), past year psychological distress (Killion et al. 2021), use of other psychoactive drugs (Killion et al. 2021; A. Yockey, King, and Vidourek 2019), previous arrests (Killion et al. 2021), risk taking behavior (A. Yockey, King, and Vidourek 2019), risk perception of LSD and other drugs (R. A. Yockey, Vidourek, and King 2020), sexually transmitted infections (STIs) (Folch et al. 2015), and history of hallucinogen abuse or dependence (Ross and Bogenschutz 2017). Based on evidence that LSD use among bisexual people may be increasing (R. A. Yockey, Vidourek, and King 2020), we included bisexual and lesbian/gay sexual identity in the models. Since mystical experiences sometimes occur in LSD users (Liechti, Dolder, and Schmid 2017), we also incorporated questions about importance of religious beliefs in the lives of survey participants. All religious factors (religious importance, religion affects decision-making, and number of religious services attended) were highly correlated, so a new variable was created that combined importance and decision-making to create a new four-level factor: religion is important, and influences my decisions, religion is important, but doesn’t influence my decisions, religion is not important, but influences my decisions, religion is not important, and doesn’t influence my decisions. This new factor was used as a representative of the respondent’s religious history.

Given the previously mentioned association between LSD use and risk-taking behavior, we incorporated two additional risk-taking variables: selling illegal drugs at least once and ever using a needle to inject drugs. LSD use has previously been associated with suicidal ideation (Han et al. 2022), so we included a past-year suicide attempt variable (except for those under 18 or in the all-ages model because respondents under 18 were not asked this question). Based on LSD use’s correlation with past arrest history, we also evaluated two related variables: ever having stolen anything worth more than $50 and ever having attacked anyone with intent to seriously hurt them. Access to psychoactive substances is associated with use (Ambrose, Cowan, and Rosenman 2021; MacNabb et al. 2022), so we incorporated variables on reported difficulty accessing LSD and whether the respondent had been approached by someone selling drugs in the previous year. Insurance status was included in the model since it is associated with some types of substance use (Tardelli et al. 2019), though we are unaware of investigations into its relationship with LSD use. Since substance use is associated with medical hospitalizations (Wu, Zhu, and Ghitza 2018), we incorporated a variable on past-year hospitalization. Because of LSD’s association with income level, we incorporated variables for current employment status, number of employers in the past year, and participation in government assistance programs in the current year (a variable combining participation in the Supplemental Security Income or “food stamps” program or receipt or either cash assistance or non-cash governmental assistance), as well as if the respondent reported living in poverty.

Working cell phone access is known to be low in some groups who use drugs (Ozga et al. 2022), but has not been explored with LSD use, so this variable was added. Finally, since parenthood is associated with decreased substance use (Fergusson, Boden, and John Horwood 2012), but also has not been investigated with LSD use, variables on the number of children under 18 years of age in the home and current pregnancy were incorporated into the models.

Factors were evaluated for collinearity, confounding, and model stability. Needle use was excluded from multivariable modeling due to low incidence. The number of employers could not be included in multivariable models, due to confounding with employment. Participation in government assistance and having a working cell phone were both not significantly associated with LSD use and wasn’t included in multivariable analysis. Due to strong correlations among all criminal variables, past arrest history was used in multivariable models as a representative factor for criminal history. Pregnancy was excluded from multivariable modeling because it was asked of female respondents only.

### Appendix B: Supplemental Tables

**Supplemental Table 1.** Drug Use Characte1istics by Past Year LSD Use from NSDUH Survey Respondents from 2015-2019.

| Survey Question | Overall | LSD Use within the past year |  |  |
| --- | --- | --- | --- | --- |
|  |  | No Past Year Use | Past Year Use |  |
| about Drug Use | (N=282,768) | (N=279,136) | (N=3,632) | p-value |
| Tobacco / Ever | 62.4 (62.0, 62.7) | 62.1 (61.8, 62.4) | 93.4 (92.4, 94.4) | $<0.001^c$ |
| Tobacco / Past Month | 22.5 (22.2, 22.8) | 22.2 (21.9, 22.5) | 62.4 (60.4, 64.4) | $<0.001^c$ |
| Tobacco / Past Year | 27.7 (27.3, 28.0) | 27.3 (27.0, 27.6) | 77.0 (74.8, 79.1) | $<0.001^c$ |
| Alcohol / Ever | 80.6 (80.4, 80.9) | 80.5 (80.2, 80.7) | 98.4 (97.9, 99.0) | $<0.001^c$ |
| Marijuana / Ever | 44.9 (44.6, 45.3) | 44.5 (44.2, 44.9) | 98.3 (97.7, 98.9) | $<0.001^c$ |
| Cocaine / Ever | 14.7 (14.5, 15.0) | 14.4 (14.1, 14.6) | 62.3 (60.2, 64.4) | $<0.001^c$ |
| "Crack" / Ever | 3.4 (3.3, 3.5) | 3.3 (3.2, 3.5) | 11.3 (9.7, 13.0) | $<0.001^c$ |
| Heroin / Ever | 1.9 (1.8, 2.0) | 1.9 (1.8, 1.9) | 12.0 (10.7, 13.3) | $<0.001^c$ |
| PCP / Ever | 2.2 (2.1, 2.3) | 2.2 (2.1, 2.3) | 6.4 (5.3, 7.5) | $<0.001^c$ |
| Ecstasy / Ever | 7.1 (6.9, 7.2) | 6.6 (6.5, 6.8) | 67.5 (65.3, 69.7) | $<0.001^c$ |
| Ketamine / Ever | 1.3 (1.2, 1.3) | 1.1 (1.07, 1.2) | 21.5 (19.7, 23.2) | $<0.001^c$ |
| Inhalants / Ever | 9.2 (9.0, 9.4) | 9.0 (8.8, 9.1) | 40.5 (38.1, 43.0) | $<0.001^c$ |
| Methamphetamine / Ever | 5.5 (5.4, 5.7) | 5.4 (5.3, 5.6) | 20.3 (18.0, 22.5) | $<0.001^c$ |
| Pain Relievers / Ever Misused | 9.8 (9.6, 10.0) | 9.5 (9.4, 9.7) | 47.8 (45.4, 50.1) | $<0.001^c$ |
| Stimulants / Ever Misused | 4.1 (4.0, 4.2) | 3.7 (3.7, 3.8) | 46.7 (44.3, 49.1) | $<0.001^c$ |
| Tranquilizers / Ever Misused | 4.3 (4.1, 4.4) | 4.0 (3.9, 4.1) | 38.2 (36.0, 40.4) | $<0.001^c$ |
| Sedative / Ever Misused | 2.2 (2.1, 2.3) | 2.2 (2.1, 2.2) | 12.9 (11.5, 14.4) | $<0.001^c$ |

**Supplemental Table 1. Drug Use Characteristics by Past Year LSD Use from NSDUH Survey Respondents from 2015-2019.**
| Survey Question | Overall | LSD Use within the past year |  |  |
| --- | --- | --- | --- | --- |
|  |  | No Past Year Use | Past Year Use |  |
| about Drug Use | (N=282,768) | (N=279,136) | (N=3,632) | p-value |
| Peyote / Ever | 2.1 (2.0, 2.2) | 2.1 (2.0, 2.2) | 7.4 (6.1, 8.7) | **<0.001<sup>c</sup>** |
| Mescaline / Ever | 2.9 (2.8, 3.0) | 2.8 (2.7, 2.9) | 10.0 (8.7, 11.3) | **<0.001<sup>c</sup>** |
| Psilocybin (mushrooms) / Ever | 8.8 (8.6, 9.0) | 8.4 (8.2, 8.6) | 68.8 (66.5, 71.2) | **<0.001<sup>c</sup>** |
| Hallucinogen Dependence or Abuse / Past Year | 0.11 (0.10, 0.13) | 0.06 (0.05, 0.07) | 6.6 (5.4, 7.9) | **<0.001<sup>c</sup>** |
| Needle Use / Ever | 1.7 (1.6, 1.7) | 1.6 (1.6, 1.7) | 7.1 (6.0, 8.3) | **<0.001<sup>c</sup>** |
| How did you get the needle last time, *Needle users only* |  |  |  | 0.07<sup>c</sup> |
| Bought/stole/given from pharm, needle exch, lab, store | 57.9 (55.7, 60.1) | 57.5 (55.2, 59.9) | 68.1 (58.7, 77.5) |  |
| Bought/stole/given from friend, relative, work, home | 24.1 (22.5, 25.7) | 24.3 (22.6, 26.0) | 17.4 (9.0, 25.7) |  |
| included with drugs | 0.88 (0.47, 1.3) | 0.91 (0.48, 1.3) | 0.09 (0.00, 0.26) |  |
| Acquired on street, found in trash can, shared needle | 8.7 (7.6, 9.7) | 8.6 (7.5, 9.7) | 9.9 (3.3, 16.6) |  |
| Some other way | 8.5 (6.9, 10.1) | 8.6 (7.0, 10.3) | 4.5 (1.3, 7.8) |  |
| How difficult to get LSD |  |  |  | **<0.001<sup>c</sup>** |
| Probably Impossible | 41.0 (40.6, 41.3) | 41.2 (40.9, 41.6) | 3.2 (2.1, 4.2) |  |
| Very Difficult | 23.3 (23.0, 23.6) | 23.4 (23.1, 23.7) | 12.3 (10.6, 14.0) |  |
| Fairly Difficult | 16.8 (16.5, 17.0) | 16.6 (16.4, 16.9) | 31.4 (29.2, 33.7) |  |
| Fairly Easy | 9.3 (9.1, 9.4) | 9.1 (8.9, 9.2) | 34.3 (31.6, 36.9) |  |
| Very Easy | 3.8 (3.7, 3.9) | 3.7 (3.6, 3.8) | 18.7 (17.0, 20.3) |  |
| Don't Know | 5.9 (5.7, 6.0) | 5.9 (5.8, 6.1) | 0.14 (0.00, 0.32) |  |
| Approached by drug seller / Past Month |  |  |  | **<0.001<sup>c</sup>** |
| Yes | 5.6 (5.5, 5.8) | 5.4 (5.3, 5.5) | 39.5 (37.6, 41.4) |  |
| No | 94.4 (94.2, 94.5) | 94.6 (94.5, 94.7) | 60.5 (58.6, 62.4) |  |

**Supplemental Table 1. Drug Use Characteristics by Past Year LSD Use from NSDUH Survey Respondents from 2015-2019.**
| Survey Question | Overall | LSD Use within the past year |  |  |
| --- | --- | --- | --- | --- |
|  |  | No Past Year Use | Past Year Use |  |
| about Drug Use | (N=282,768) | (N=279,136) | (N=3,632) | p-value |
| Frequencies presented are unweighted counts. Weighted proportions and 95% confidence intervals are reported. *Imputed or recoded variable reported. P-values: a=linear regression; b=linear regression with log transformation; c=Rao-Scott chi-square test. SAS Survey Procedures used for all analyses. |  |  |  |  |

**Supplemental Table 2.** Risk Attitudes by Past-Year LSD Use from NSDUH Survey Respondents from 2015-2019.

| Survey Question | Overall | LSD Use within the past year |  |  |
| --- | --- | --- | --- | --- |
|  |  | No Past Year Use | Past Year Use |  |
|  | (N=282,768) | (N=279,136) | (N=3,632) | p-value |
| Risk Smoking Marijuana Once or Twice A Week |  |  |  | **<0.001<sup>c</sup>** |
| No Risk | 18.9 (18.6, 19.1) | 18.5 (18.2, 18.8) | 65.9 (63.9, 68.0) |  |
| Slight Risk | 24.6 (24.4, 24.9) | 24.6 (24.3, 24.9) | 26.9 (25.0, 28.7) |  |
| Moderate Risk | 23.1 (22.8, 23.4) | 23.3 (23.0, 23.5) | 5.7 (4.5, 7.0) |  |
| Great Risk | 31.8 (31.5, 32.1) | 32.0 (31.7, 32.3) | 1.4 (0.86, 1.9) |  |
| Don't Know | 1.6 (1.5, 1.7) | 1.6 (1.5, 1.7) | 0.11 (0.00, 0.23) |  |
| Risk Trying LSD Once or Twice |  |  |  | **<0.001<sup>c</sup>** |
| No Risk | 3.5 (3.4, 3.6) | 3.2 (3.1, 3.3) | 34.9 (32.5, 37.3) |  |
| Slight Risk | 10.9 (10.7, 11.1) | 10.7 (10.5, 10.9) | 43.4 (41.2, 45.6) |  |
| Moderate Risk | 16.8 (16.6, 17.0) | 16.8 (16.6, 17.0) | 14.1 (12.5, 15.7) |  |
| Great Risk | 66.3 (66.0, 66.7) | 66.8 (66.4, 67.1) | 7.4 (6.3, 8.5) |  |
| Don't Know | 2.5 (2.4, 2.6) | 2.5 (2.4, 2.6) | 0.22 (0.00, 0.50) |  |
| Risk Using LSD Once or Twice A Week |  |  |  | **<0.001<sup>c</sup>** |

| Survey Question | Overall<br>(N=282,768) | LSD Use within the past year |  |  |
| --- | --- | --- | --- | --- |
|  |  | No Past Year Use<br>(N=279,136) | Past Year Use<br>(N=3,632) | p-value |
| No Risk | 1.5 (1.4, 1.5) | 1.4 (1.3, 1.4) | 12.0 (10.6, 13.5) |  |
| Slight Risk | 3.5 (3.4, 3.7) | 3.4 (3.3, 3.5) | 23.0 (21.1, 24.9) |  |
| Moderate Risk | 12.1 (12.0, 12.3) | 12.0 (11.8, 12.2) | 32.2 (30.1, 34.2) |  |
| Great Risk | 80.4 (80.1, 80.6) | 80.7 (80.5, 81.0) | 32.6 (30.0, 35.1) |  |
| Don't Know | 2.5 (2.4, 2.6) | 2.5 (2.4, 2.6) | 0.23 (0.02, 0.44) |  |
| Risk Trying Heroin Once or Twice |  |  |  | **<0.001<sup>c</sup>** |
| No Risk | 1.2 (1.1, 1.2) | 1.2 (1.1, 1.2) | 2.3 (1.5, 3.0) |  |
| Slight Risk | 3.6 (3.4, 3.7) | 3.5 (3.4, 3.7) | 7.1 (5.5, 8.7) |  |
| Moderate Risk | 9.3 (9.1, 9.5) | 9.3 (9.1, 9.5) | 15.1 (13.1, 17.1) |  |
| Great Risk | 84.3 (84.0, 84.6) | 84.4 (84.1, 84.6) | 75.1 (72.4, 77.7) |  |
| Don't Know | 1.6 (1.5, 1.7) | 1.6 (1.5, 1.7) | 0.48 (0.17, 0.79) |  |
| Risk Using Heroin Once or Twice A Week |  |  |  | **0.002<sup>c</sup>** |
| No Risk | 0.91 (0.86, 0.96) | 0.91 (0.86, 0.96) | 0.83 (0.37, 1.3) |  |
| Slight Risk | 0.99 (0.94, 1.05) | 0.99 (0.94, 1.05) | 1.1 (0.54, 1.7) |  |
| Moderate Risk | 3.9 (3.7, 4.0) | 3.9 (3.7, 4.0) | 4.8 (3.8, 5.8) |  |
| Great Risk | 92.7 (92.5, 92.8) | 92.7 (92.5, 92.8) | 92.8 (91.5, 94.1) |  |
| Don't Know | 1.6 (1.5, 1.6) | 1.6 (1.5, 1.6) | 0.44 (0.15, 0.73) |  |
| Risk Using Cocaine Once or Twice A Week |  |  |  | **<0.001<sup>c</sup>** |
| No Risk | 1.04 (0.99, 1.09) | 1.04 (0.99, 1.09) | 1.3 (0.79, 1.8) |  |
| Slight Risk | 2.6 (2.4, 2.7) | 2.5 (2.4, 2.6) | 10.8 (9.1, 12.4) |  |

|  | Overall | LSD Use within the past year |  |  |
| --- | --- | --- | --- | --- |
|  |  | No Past Year Use | Past Year Use |  |
| Survey Question | (N=282,768) | (N=279,136) | (N=3,632) | p-value |
| Moderate Risk | 9.5 (9.3, 9.7) | 9.4 (9.2, 9.6) | 30.7 (28.1, 33.2) |  |
| Great Risk | 85.3 (85.1, 85.6) | 85.5 (85.3, 85.8) | 57.1 (54.7, 59.5) |  |
| Don't Know | 1.6 (1.5, 1.6) | 1.6 (1.5, 1.7) | 0.17 (0.04, 0.29) |  |
| Risk Having 5 or More Drinks Once or Twice A Week |  |  |  | **<0.001<sup>c</sup>** |
| No Risk | 3.2 (3.1, 3.3) | 3.1 (3.0, 3.2) | 5.8 (4.6, 7.0) |  |
| Slight Risk | 16.6 (16.4, 16.8) | 16.5 (16.3, 16.7) | 30.7 (28.0, 33.4) |  |
| Moderate Risk | 35.1 (34.9, 35.4) | 35.1 (34.8, 35.4) | 39.0 (36.2, 41.8) |  |
| Great Risk | 44.2 (43.9, 44.5) | 44.4 (44.1, 44.7) | 24.3 (22.3, 26.3) |  |
| Don't Know | 0.91 (0.85, 0.96) | 0.91 (0.86, 0.97) | 0.18 (0.04, 0.33) |  |
| Get A Real Kick Out Of Doing Dangerous Things |  |  |  | **<0.001<sup>c</sup>** |
| Never | 54.3 (53.9, 54.6) | 54.6 (54.3, 54.9) | 10.5 (9.0, 12.0) |  |
| Seldom | 28.2 (27.9, 28.5) | 28.2 (27.9, 28.5) | 27.8 (25.6, 30.0) |  |
| Sometimes | 15.0 (14.8, 15.2) | 14.7 (14.5, 14.9) | 47.5 (45.4, 49.5) |  |
| Always | 2.6 (2.5, 2.7) | 2.5 (2.4, 2.6) | 14.2 (12.7, 15.6) |  |
| Like To Test Yourself By Doing Risky Things |  |  |  | **<0.001<sup>c</sup>** |
| Never | 53.9 (53.6, 54.2) | 54.2 (53.9, 54.5) | 13.7 (12.1, 15.2) |  |
| Seldom | 32.0 (31.7, 32.3) | 32.0 (31.7, 32.3) | 37.4 (35.1, 39.7) |  |
| Sometimes | 12.5 (12.4, 12.7) | 12.3 (12.2, 12.5) | 39.8 (37.8, 41.9) |  |
| Always | 1.6 (1.5, 1.7) | 1.5 (1.5, 1.6) | 9.1 (8.0, 10.2) |  |

|  | Overall | LSD Use within the past year |  |  |
| --- | --- | --- | --- | --- |
|  |  | No Past Year Use | Past Year Use |  |
| Survey Question | (N=282,768) | (N=279,136) | (N=3,632) | p-value |

**Supplemental Table 3.** Drug Use Characteristics for all Study Respondents by Past-Year LSD Use per Study Year.

|  | 2015 | 2016 | 2017 | 2018 | 2019 | 2015<br>vs<br>2019 | Year<br>x<br>Var |
| --- | --- | --- | --- | --- | --- | --- | --- |
| Demographic Characteristics | Statistics | (N=56,897) | (N=56,276) | (N=56,313) | (N=56,136) | p-value | p-value |
| Past Year LSD Use | 15.2<br>(13.7,<br>16.7) | 18.4<br>(16.6,<br>20.2) | 21.2<br>(19.3,<br>23.0) | 22.2<br>(20.2,<br>24.2) | 23.1<br>(21.1,<br>25.1) | <.0001 | <.0001 |
| ANY TOBACCO - EVER USED |  |  |  |  |  |  |  |
| No LSD Use | 63.6<br>(62.9,<br>64.2) | 62.7<br>(62.1,<br>63.4) | 62.4<br>(61.8,<br>63.0) | 61.2<br>(60.5,<br>61.9) | 60.8<br>(60.0,<br>61.6) | <.0001 | 0.35 |
| Past Year LSD Use | 93.7<br>(91.5,<br>95.9) | 93.3<br>(90.6,<br>96.0) | 93.8<br>(91.6,<br>96.0) | 94.8<br>(93.0,<br>96.7) | 91.7<br>(88.8,<br>94.5) |  |  |
| ANY TOBACCO - PAST YEAR USE |  |  |  |  |  |  |  |
| No LSD Use | 29.1<br>(28.5,<br>29.7) | 28.2<br>(27.6,<br>28.8) | 27.0<br>(26.4,<br>27.6) | 26.3<br>(25.6,<br>27.0) | 25.9<br>(25.3,<br>26.5) | <.0001 | 0.55 |
| Past Year LSD Use | 82.8<br>(79.4,<br>86.2) | 77.5<br>(73.1,<br>81.8) | 77.8<br>(72.8,<br>82.8) | 75.9<br>(71.3,<br>80.6) | 72.9<br>(67.5,<br>78.3) |  |  |
| ALCOHOL - EVER USED |  |  |  |  |  |  |  |
| No LSD Use | 80.9<br>(80.4,<br>81.4) | 80.0<br>(79.6,<br>80.5) | 80.8<br>(80.3,<br>81.3) | 80.6<br>(80.1,<br>81.1) | 80.1<br>(79.5,<br>80.7) | 0.06 | 0.77 |
| Past Year LSD Use | 98.1<br>(96.9,<br>99.3) | 98.5<br>(97.4,<br>99.7) | 98.5<br>(97.5,<br>99.5) | 98.2<br>(97.1,<br>99.3) | 98.7<br>(97.6,<br>99.8) |  |  |
| MARIJUANA -<br>EVER USED |  |  |  |  |  |  |  |
| No LSD Use | 43.7<br>(42.8,<br>44.5) | 43.5<br>(42.8,<br>44.2) | 44.7<br>(43.9,<br>45.4) | 44.8<br>(44.0,<br>45.6) | 45.9<br>(45.2,<br>46.7) | **<.00**<br>**01** | 0.18 |
| Past Year LSD Use | 97.1<br>(95.3,<br>99.0) | 97.7<br>(95.6,<br>99.7) | 98.8<br>(97.8,<br>99.8) | 99.0<br>(98.3,<br>99.8) | 98.3<br>(97.3,<br>99.3) |  |  |
| COCAINE - EVER<br>USED |  |  |  |  |  |  |  |
| No LSD Use | 14.3<br>(13.8,<br>14.8) | 14.1<br>(13.6,<br>14.6) | 14.5<br>(14.1,<br>14.9) | 14.3<br>(13.8,<br>14.9) | 14.6<br>(14.1,<br>15.1) | 0.25 | 0.19 |
| Past Year LSD Use | 60.3<br>(55.9,<br>64.6) | 58.0<br>(53.3,<br>62.7) | 64.1<br>(59.9,<br>68.2) | 64.0<br>(59.2,<br>68.8) | 63.8<br>(59.4,<br>68.2) |  |  |
| "CRACK" - EVER<br>USED |  |  |  |  |  |  |  |
| No LSD Use | 3.3<br>(3.0,<br>3.5) | 3.2 (3.0,<br>3.4) | 3.5 (3.2,<br>3.7) | 3.4 (3.2,<br>3.7) | 3.3 (3.1,<br>3.6) | 0.51 | 0.17 |
| Past Year LSD Use | 10.4<br>(6.9,<br>13.9) | 7.9 (4.9,<br>10.8) | 13.0<br>(9.5,<br>16.4) | 11.5<br>(7.3,<br>15.7) | 13.1<br>(9.9,<br>16.2) |  |  |
| HEROIN - EVER<br>USED |  |  |  |  |  |  |  |
| No LSD Use | 1.8<br>(1.6,<br>2.0) | 1.7 (1.6,<br>1.9) | 1.9 (1.7,<br>2.1) | 1.8 (1.7,<br>2.0) | 2.0 (1.8,<br>2.2) | 0.17 | 0.79 |
| Past Year LSD Use | 13.9<br>(10.1,<br>17.7) | 12.1<br>(9.0,<br>15.1) | 13.7<br>(10.1,<br>17.3) | 11.8<br>(8.7,<br>14.9) | 9.4 (6.4,<br>12.4) |  |  |
| PCP - EVER USED |  |  |  |  |  |  |  |
| No LSD Use | 2.3<br>(2.1,<br>2.5) | 2.3 (2.1,<br>2.5) | 2.3 (2.1,<br>2.5) | 2.3 (2.0,<br>2.5) | 2.0 (1.7,<br>2.2) | **0.04** | 0.37 |
| Past Year LSD Use | 8.6<br>(4.9,<br>12.4) | 7.7 (5.0,<br>10.3) | 5.3 (3.2,<br>7.5) | 4.9 (2.9,<br>6.9) | 6.4 (4.0,<br>8.8) |  |  |
| ECSTASY - EVER USED |  |  |  |  |  |  |  |
| No LSD Use | 6.6<br>(6.3, 6.9) | 6.3 (6.1, 6.6) | 6.6 (6.3, 6.9) | 6.9 (6.5, 7.2) | 6.7 (6.4, 7.0) | 0.11 | 0.10 |
| Past Year LSD Use | 74.3<br>(69.6, 78.9) | 64.3<br>(59.7, 68.8) | 63.3<br>(57.5, 69.0) | 69.2<br>(65.2, 73.3) | 67.8<br>(63.3, 72.3) |  |  |
| KETAMINE - EVER USED |  |  |  |  |  |  |  |
| No LSD Use | 1.05<br>(0.94, 1.2) | 1.1<br>(1.00, 1.2) | 1.1<br>(1.01, 1.2) | 1.2<br>(1.01, 1.3) | 1.2<br>(1.04, 1.3) | **0.01** | 0.47 |
| Past Year LSD Use | 20.5<br>(15.2, 25.9) | 20.9<br>(16.4, 25.5) | 20.3<br>(15.5, 25.1) | 20.0<br>(15.9, 24.2) | 25.0<br>(20.0, 29.9) |  |  |
| INHALANTS - EVER USED |  |  |  |  |  |  |  |
| No LSD Use | 9.3<br>(9.0, 9.7) | 8.9 (8.5, 9.2) | 9.1 (8.7, 9.5) | 8.8 (8.4, 9.2) | 8.8 (8.4, 9.2) | 0.06 | 0.08 |
| Past Year LSD Use | 41.3<br>(36.2, 46.4) | 36.9<br>(31.6, 42.2) | 38.4<br>(34.3, 42.5) | 45.6<br>(40.8, 50.4) | 40.0<br>(34.8, 45.2) |  |  |
| METHAMPHETAMINE - EVER USED |  |  |  |  |  |  |  |
| No LSD Use | 5.4<br>(5.1, 5.7) | 5.3 (5.1, 5.5) | 5.3 (5.0, 5.7) | 5.3 (5.1, 5.6) | 5.7 (5.3, 6.1) | 0.12 | **0.04** |
| Past Year LSD Use | 14.0<br>(9.9, 18.2) | 19.5<br>(15.3, 23.7) | 22.9<br>(18.4, 27.5) | 20.2<br>(15.5, 24.9) | 22.6<br>(18.6, 26.6) |  |  |
| PAIN RELIEVERS - EVER MISUSED |  |  |  |  |  |  |  |
| No LSD Use | 10.1<br>(9.7, 10.4) | 9.8 (9.5, 10.2) | 9.6 (9.2, 10.0) | 9.1 (8.7, 9.5) | 9.1 (8.7, 9.4) | **0.0002** | 0.95 |
| Past Year LSD Use | 51.1<br>(45.6, 56.7) | 48.9<br>(42.2, 55.5) | 49.6<br>(45.3, 53.9) | 45.5<br>(40.7, 50.2) | 45.2<br>(40.3, 50.2) |  |  |
| STIMULANTS - EVER MISUSED |  |  |  |  |  |  |  |
| No LSD Use | 3.5<br>(3.3, 3.7) | 3.9 (3.7, 4.2) | 3.9 (3.7, 4.1) | 3.5 (3.3, 3.7) | 3.8 (3.6, 4.0) | **0.01** | 0.87 |
| Past Year LSD Use | 49.4<br>(45.1,<br>53.8) | 47.2<br>(41.7,<br>52.8) | 47.2<br>(42.1,<br>52.3) | 45.4<br>(41.3,<br>49.5) | 45.2<br>(39.6,<br>50.7) |  |  |
| TRANQUILIZERS -<br>EVER MISUSED |  |  |  |  |  |  |  |
| No LSD Use | 4.0<br>(3.8,<br>4.3) | 4.2 (3.9,<br>4.4) | 4.1 (3.8,<br>4.3) | 3.9 (3.6,<br>4.1) | 3.9 (3.6,<br>4.1) | 0.65 | 0.65 |
| Past Year LSD Use | 38.0<br>(32.6,<br>43.4) | 40.3<br>(36.3,<br>44.3) | 37.6<br>(32.3,<br>42.9) | 38.9<br>(34.6,<br>43.1) | 36.6<br>(31.2,<br>42.0) |  |  |
| SEDATIVES - EVER<br>MISUSED |  |  |  |  |  |  |  |
| No LSD Use | 2.1<br>(1.9,<br>2.2) | 2.3 (2.1,<br>2.5) | 2.3 (2.1,<br>2.4) | 2.0 (1.8,<br>2.2) | 2.1 (1.9,<br>2.3) | 0.68 | 0.87 |
| Past Year LSD Use | 13.7<br>(10.5,<br>17.0) | 12.9<br>(9.2,<br>16.6) | 12.8<br>(9.5,<br>16.1) | 13.4<br>(9.9,<br>16.9) | 12.1<br>(8.4,<br>15.8) |  |  |
| EVER USED<br>PEYOTE |  |  |  |  |  |  |  |
| No LSD Use | 2.0<br>(1.8,<br>2.3) | 2.1 (1.9,<br>2.3) | 2.0 (1.8,<br>2.2) | 2.2 (2.0,<br>2.4) | 2.0 (1.8,<br>2.3) | 1.00 | 0.43 |
| Past Year LSD Use | 8.4<br>(5.2,<br>11.5) | 8.4 (5.7,<br>11.2) | 5.1 (2.7,<br>7.5) | 9.3 (6.1,<br>12.5) | 6.3 (3.9,<br>8.7) |  |  |
| EVER USED<br>MESCALINE |  |  |  |  |  |  |  |
| No LSD Use | 3.0<br>(2.6,<br>3.3) | 3.0 (2.8,<br>3.2) | 2.7 (2.5,<br>3.0) | 2.7 (2.5,<br>3.0) | 2.7 (2.4,<br>2.9) | 0.19 | 0.81 |
| Past Year LSD Use | 12.5<br>(9.0,<br>16.0) | 10.5<br>(6.8,<br>14.1) | 9.5 (6.9,<br>12.1) | 8.7 (4.8,<br>12.5) | 9.7 (6.8,<br>12.7) |  |  |
| EVER USED<br>PSILOCYBIN<br>(MUSHROOMS) |  |  |  |  |  |  |  |
| No LSD Use | 8.2<br>(7.8,<br>8.6) | 8.0 (7.6,<br>8.3) | 8.4 (8.0,<br>8.7) | 8.8 (8.4,<br>9.2) | 8.6 (8.1,<br>9.0) | **0.046** | 0.14 |
| Past Year LSD Use | 68.8<br>(63.9,<br>73.6) | 64.0<br>(58.3,<br>69.7) | 66.9<br>(62.2,<br>71.5) | 71.8<br>(67.1,<br>76.5) | 71.6<br>(67.8,<br>75.5) |  |  |
| HALLUCINOGEN<br>DEPENDENCE OR |  |  |  |  |  |  |  |
| ABUSE - PAST YEAR |  |  |  |  |  |  |  |
| No LSD Use | 0.05<br>(0.04, 0.07) | 0.08<br>(0.06, 0.11) | 0.07<br>(0.04, 0.09) | 0.05<br>(0.03, 0.07) | 0.06<br>(0.04, 0.09) | 0.81 | 0.42 |
| Past Year LSD Use | 7.9<br>(5.1, 10.6) | 7.3 (4.7, 10.0) | 7.1 (4.7, 9.5) | 6.6 (4.8, 8.4) | 4.8 (2.9, 6.8) |  |  |
| EVER USED NEEDLE TO INJECT DRUGS |  |  |  |  |  |  |  |
| No LSD Use | 1.6<br>(1.4, 1.8) | 1.6 (1.4, 1.8) | 1.6 (1.4, 1.8) | 1.6 (1.4, 1.8) | 1.7 (1.5, 1.8) | 0.67 | 0.94 |
| Past Year LSD Use | 7.3<br>(4.7, 9.9) | 7.9 (5.0, 10.8) | 7.2 (4.6, 9.8) | 6.7 (4.4, 9.1) | 6.7 (4.1, 9.4) |  |  |
| SOLD ILLEGAL DRUGS |  |  |  |  |  |  |  |
| No LSD Use | 1.4<br>(1.3, 1.5) | 1.4 (1.3, 1.6) | 1.4 (1.2, 1.5) | 1.3 (1.2, 1.5) | 1.3 (1.1, 1.4) | 0.39 | 0.99 |
| Past Year LSD Use | 25.1<br>(20.8, 29.5) | 24.5<br>(19.8, 29.1) | 23.9<br>(19.6, 28.2) | 22.0<br>(17.5, 26.4) | 21.6<br>(17.9, 25.4) |  |  |
| All survey questions on drug use were recoded based on previous survey responses. Frequencies presented are unweighted counts. All weighted frequencies are reported as % (95% CI). All associations with Lifetime LSD Use within each survey year were significant, $p < 0.001$ (Rao-Scott chi-square test). All associations are adjusted for survey year, age groups, gender, race, employment status, marital status, education, religious beliefs, criminal arrest, and county metro area. SAS Survey Procedures used for all analyses. |  |  |  |  |  |  |  |

